# Defective B cell tolerance in SLE lymph nodes underpins VH_4-34_ “clonal damnation” and PD-1⁺TOX⁺ autoreactive B cells expansion

**DOI:** 10.64898/2026.05.18.26353148

**Authors:** Caterina E. Faliti, Midushi Ghimire, Melissa Garcia-Vega, Rachel C. Watermeier, Amanda Callahan, Julia Burke, Olivia Posadas, Ashish K. Mishra, Surender Khurana, Victor Greiff, Chris D. Scharer, John M. Lindner, R. Glenn King, Mary Newell, Arezou Khosroshahi, F. Eun-Hyung Lee, Iñaki Sanz

**Author notes:** Corresponding authors: I.S., C.E.F.

## Abstract

Systemic Lupus Erythematosus (SLE) is a chronic autoimmune disease driven by uncensored B and T cell autoreactivity. Understanding this pathogenic process has been hampered by lack of studies of secondary lymphoid organs in human SLE. Using minimally invasive lymph node fine needle aspirates (LN-FNAs), we profiled tissue-resident immune cells from 59 SLE patients and 34 healthy controls through high-dimensional 43-color flow cytometry, antigen-specific tetramer probing, and sc-RNA sequencing with paired VH/VL repertoire analysis. Our findings reveal hyperactive lymph node immunity in SLE characterized by spontaneous germinal center (GC) activation, plasma cell accumulation enriched in mature CD19^-^ and CD138^+^ antibody-secreting cells, and increased frequencies of both GC-T_FH_ and PD-1^+^CXCR5^-^ T extra-follicular helper cells. SLE lymph nodes harbored large oligoclonal B cell families with altered isotype usage, dominated by IgG1 and IgG4. Critically, self-reactive 9G4^+^ and Ro60^+^ B cells showed defective tolerance checkpoint control, accumulating in activated naïve, GC, and plasma cell compartments with distinctive PD-1^+^Tox^+^ expression absent in viral-specific responses. Single-cell repertoire analysis revealed VH_4-34_ clones in SLE B_GC_ and B_PC_, that in contrast to HD, had not experienced clonal redemption. Instead, SLE VH_4-34_ clones displayed low somatic hypermutation and preserved the AVY hydrophobic patch associated with autoreactivity. Monoclonal antibody testing confirmed that unmutated AVY^+^ VH_4-34_ clones retained polyreactivity against naïve B cells, apoptotic cells, and multiple self-antigens. Together, these results define “clonal damnation” as a key mechanism in SLE whereby autoreactive VH_4-34_ clones of pathogenic potential escape tolerance checkpoints, expand in germinal centers, and differentiate into tissue plasma cells while preserving germline-encoded self-reactivity. Combined, our study defines critical mechanisms of tolerance breakdown in lupus pathogenesis.

## Introduction

Systemic Lupus Erythematosus (SLE) is a prototypic autoimmune disease characterized by failure of immune tolerance and the expansion of self-reactive lymphocyte clones with pathogenic potential^1, 2, 3, 4^. A central role for pathogenic B cells is supported by the clinical efficacy of B cell depletion, more recently including cellular therapies that can induce immune resetting and prolonged drug-free remission^5, 6, 7^. Yet, the mechanisms that regulate human B cell differentiation and enforce tolerance, remain poorly understood, in large part due to lack of studies of secondary lymphoid tissues, where germinal center (GC) reactions and selection of autoreactive clones occur. We have addressed these limitations by studying human lymph node B cell responses using ultra-sound guided fine-needle aspirates of axillary lymph nodes (LN-FNAs). In this study, we analyzed autoreactive B cells expressing the 9G4 idiotype and anti-Ro60 B cells in LNs of SLE patients and Healthy Donors (HD). The 9G4 idiotype is determined by the expression of VH_4-34_-encoded antibodies with preservation of a germline encoded framework-1 (FR1), hydrophobic patch (AVY). This FR1 sequence imparts intrinsic autoreactivity to 9G4^+^ antibodies, which recognize n-acetyl-lactosamine glycans shared by Ii blood group antigens and B cells, thereby determining the agglutinin properties of IgM 9G4 antibodies and the anti-B cell activity of IgG 9G4 antibodies in SLE. Owing to these properties, the expression of the 9G4 idiotype is virtually synonymous with the presence of autoreactivity of pathogenic potential. Indeed, elevated levels of serum 9G4 IgG antibodies are highly specific for SLE, correlate with anti-dsDNA antibodies and contribute a large fraction of this pathogenic autoreactivity^8, 9^ as the antibodies against DNAse1L3, highly correlated with lupus nephritis. In turn, Ro60 autoantibodies are present in large amounts in the serum of ∼35-70% of lupus patients and may contribute to disease in different ways, including the formation of immune complexes and the induction of type I interferon secretion. A pathogenic role is also suggested by their association with disease manifestations such as photosensitive rash, vasculitis and hypocomplementemia^10^. Of significant interest, these autoantibodies can be detected several years before disease onset^11^ and may cross-react with the Epstein Barr Virus (EBV) antigen EBNA^12^. Under healthy conditions, autoreactive VH_4-34_ clones are regulated in the GC, where they undergo deletion or redemption, a mechanism mediated by somatic hypermutation leading to the elimination of the FR1-dependent autoreactivity^13^. While we have previously shown defective GC censoring of 9G4^+^ cells by deletion^4^, whether redemption is also faulty in SLE remains to be investigated. Also poorly understood are the fate and antigenic selection of VH_4-34_ clones that progress through the GC reaction. Similarly, little is known about the regulation of anti-Ro60 B cell responses in HD or their dysregulation in SLE, generally believed to induce long-lived bone plasma cells responsible for high levels of serum autoantibodies that are insensitive to Lupus therapies, including B cell depletion induced by CD19 CAR-T^5, 6^.

Here, we sought to answer these questions through the combined analysis of cellular profiles, B cell repertoire and antigenic reactivity of human LN in SLE and HD. Using this approach, we have generated the first *in-depth* characterization of tissue-based B cell and T cell immune responses in human SLE with particular emphasis on the evolution of autoreactive clones within germinal centers and their differentiation into antibody-secreting plasma cells (PC).

### Study design and cohort’ characterization

In this study, we explore the regulation of B cells in secondary lymphoid tissues sampled by Fine-Needle Aspirate (FNA) via ultrasound-guided biopsies of human axillary Lymph Nodes (LNs) (**Fig. 1a**). We enrolled adults (18-65 years old, median 37 years old), age- and gender-matched, from two cohorts, Healthy Donors (HD, n=34) and patients diagnosed with Systemic Lupus Erythematosus (SLE, n=59). The main characteristics of these two cohorts are reported in **Supplemental Table 1**. LN-FNA biopsies provided an average of ∼4×10^6^ viable cells (**Extended Data Fig. 1a**), that enable deep phenotypic profiling of lymph node B cells, plasma cells and other immune populations. LN-FNA samples were analyzed using high-dimensional flow-cytometry on n=54 and n=79 samples collected from HD and SLE, respectively (**Fig. 1b**). The enrolled subjects were also characterized for their seroreactivities against various viral- and self-antigens. Furthermore, scRNAseq transcriptomics and repertoire analyses from two independent experiments using the high-throughput single-cell 10X Genomics platform allowed us to characterize the lymph node BCR repertoire from a total of six HD and eight SLE selected subjects (**Extended Data Fig. 1b, c**). Given the clinical limitations imposed by more severe disease, the majority of the enrolled SLE patients (∼73%) had a low disease activity score (SELENA-SLEDAI score of ≤4) (**Extended Data Fig. 1d**).

**Fig. 1.**
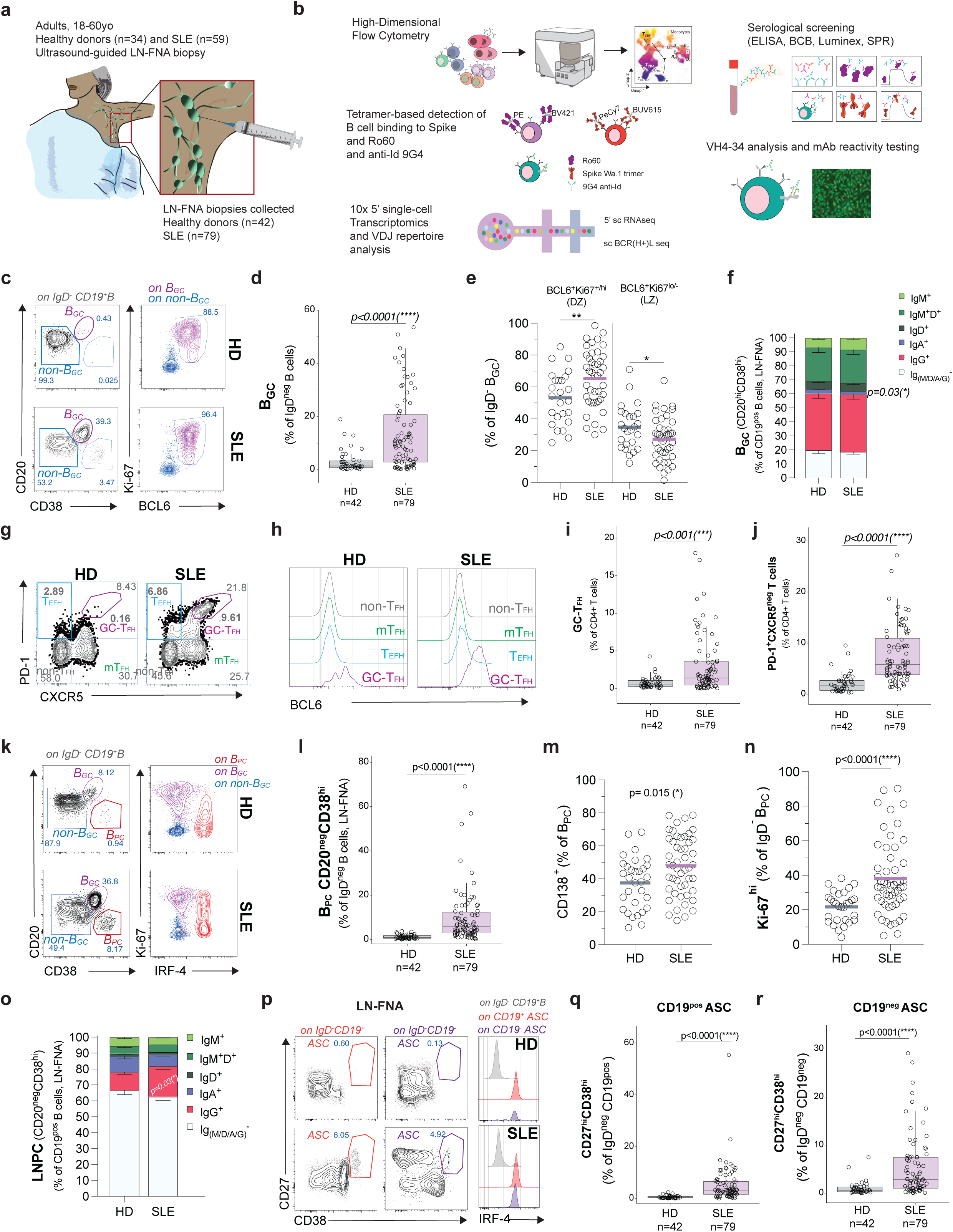
Characterization of human SLE lymph nodes. **a,** Cartoon showing the donors’ enrollment characteristics for the LN-FNA biopsy collection. **b,** Schematics showing the system-level immunoprofiling of lymph node resident immune cells and B cell reactivity tested in the study. **c,** Representative flow plots showing the gating strategy for germinal center B cells (B_GC_) in human lymph nodes. B_GC_ are identified as CD38^hi^CD20^hi^B cells among IgD^-^ CD19^+^ B cells. The flow plots show also the expression of intracellular Ki-67 and BCL6 markers gated on both B_GC_ (violet) and non-B_GC_ (light blue, CD38^-^CD20^+^). **d,** Bar plots showing the quantitation of B_GC_ in healthy donors (HD, n=42) and lupus (SLE, n=79) patients. Horizontal lines are median. Each dot represents an individual sample. Mann Whitney U test shown for comparisons. **e,** Scatter plots showing the percentage of DZ-like (BCL6^+^Ki67^+/hi^) and LZ-like (BCL6^+^Ki67^lo/-^) B_GC_ cells. Each dot represents an individual sample. Mann Whitney U test shown for comparisons. **f,** Bar plots showing the distribution of Ig isotypes among total B_GC_ gated as CD19^+^CD38^hi^CD20^hi^ B cells. Bars represent Standard Error Mean (S.E.M). Mann Whitney U test shown for comparisons between HD and SLE isotype class. **g,** Representative flow plots showing the gating strategy for germinal center T helper cells (GC-T_FH_) and extra-follicular T helper cells (T_EFH_) in human lymph nodes. GC-T_FH_ are identified as CXCR5^hi^PD-1^hi^ T cells among CD3^+^CD4^+^ T cells; T_EFH_ are identified as CXCR5^-^PD-1^hi^ T cells among CD3^+^CD4^+^ T cells. **h,** Histograms showing the expression of intracellular BCL6 gated on four CD4^+^ T cell subsets defined by the expression of lack of CXCR5 and PD-1 (grey non-T_FH_, CXCR5^-^PD-1^-^; green mT_FH_ CXCR5^+^PD-1^-^; purple GC-T_FH_ CXCR5^hi^PD-1^hi^; light blue T_EFH_, CXCR5^-^PD-1^hi^). **i,** Bar plots showing the quantitation of GC-T_FH_ in healthy donors (HD, n=42) and lupus (SLE, n=79) patients. Horizontal lines are median. Each dot represents an individual sample. Mann Whitney U test shown for comparisons. **j,** Bar plots showing the quantitation of T_EFH_ in healthy donors (HD, n=42) and lupus (SLE, n=79) patients. Horizontal lines are median. Each dot represents an individual sample. Mann Whitney U test shown for comparisons. **k,** Representative flow plots showing the gating strategy for plasma cells (B_PC_) in human lymph nodes. B_PC_ are identified as CD38^hi^CD20^lo/-^ B cells among IgD^-^CD19^+^ B cells. The flow plots show also the expression of intracellular Ki-67 and IRF4 markers gated on B_PC_ (orange), B_GC_ (purple) and non-B_GC_ (blue). **l,** Bar plots showing the quantitation of B_PC_ in healthy donors (HD, n=42) and lupus (SLE, n=79) patients. Horizontal lines are median. Each dot represents an individual sample. Mann Whitney U test shown for comparisons. **m,** Scatter plots showing the percentage of **m**, CD138^+^ cells and **n**, Ki67^+^ cells gated on IgD^-^CD19^+^CD38^hi^CD20^lo/-^ B_PC_ cells. Each dot represents an individual sample. Mann Whitney U test shown for comparisons. **o**, Bar plots showing the distribution of Ig isotypes among total B_PC_ gated as CD19^+^CD38^hi^CD20^lo/-^ B cells. Bars represent Standard Error Mean (S.E.M). Mann Whitney U test shown for comparisons between HD and SLE isotype class. **p**, Representative flow plots showing the gating strategy for total ASC in human lymph nodes gated as CD38^hi^CD27^hi^ cells from CD19^+^ and CD19^-^ compartments. Intracellular IRF4 expression is shown to compare total IgD^-^ B cells (grey), CD19^+^ ASC (orange) and CD19^-^ ASC (purple). Bar plots showing the quantitation of ASC in healthy donors (HD, n=42) and lupus (SLE, n=79) patients for **q**, CD19^+^ ASC and **r**, CD19^-^ ASC. Horizontal lines are median. Each dot represents an individual sample. Mann Whitney U test shown for comparisons.

### Seropositivity and affinity measurements of circulating antibodies against self- and viral-antigens

SLE patients were classified on the basis of serological status for 9G4 and anti-Ro60 autoantibodies ^14, 15^ ^8, 9^ (**Supplemental Table 1**). In SLE, 72.2% and 53.2% of the tested plasma samples were positive for Ro60 and 9G4 IgG serum antibodies, respectively (**Extended Data Fig. 2a, b**). Anti-Spike, and anti-RBD antibody responses were also tested to evaluate and compare foreign, anti-viral B cell reactivity using SARS-CoV-2 a Wuhan.1 or various Omicron proteins (**Extended Data Fig. 2c).** All enrolled donors showed some level of anti-Spike/RBD IgG responses, either from their history of repeated vaccinations and/or COVID-19 infections, although none of them had received vaccinations within at least three months from the study visit, nor had any reported symptoms of infection upon enrollment. Lack of recent infection was also supported by relatively low anti-Nucleocapsid IgG titers (positivity >10^4^, **Extended Data Fig. 2d).** Antibody affinity evaluation with Surface-Plasmon Resonance (SPR) testing of sera samples revealed a positive correlation between higher IgG titers and affinity for both Spike and RBD proteins, in both cohorts. Relative to HD (**Extended Data Fig. 2e**), SLE samples had a broader range of reactivity with overall reduced antibody affinity (**Extended Data Fig. 2f**), a finding consistent with our previous report of reduced affinity maturation upon mRNA COVID-19 vaccinations^16^. Similarly, SPR testing failed to demonstrate detectable Ro60 binding in any HD samples tested. As expected, SPR analysis readily detected Ro60 binding of SLE serum samples classified as seropositive using a luciferase immunoprecipitation assay (LIP), with a narrower range of affinity across a wide range of LIP anti-Ro60 antibody titers (**Extended Data Fig. 2g**).

### Human SLE LNs harbor spontaneous germinal centers

In order to characterize the immune landscape of SLE LNs, we first quantitated the frequency of germinal-center B cells (B_GC_) in LN-FNA. B_GC_ were gated as CD19^+^IgD^-^CD20^+^CD38^+^ cells, expressing high levels of Ki-67 and BCL6 (**Fig. 1c**). While HD LNs had low-to-absent frequency of B_GC_, SLE patients displayed a significantly greater frequency of B_GC_ (HD range 0.25-19%, median 5.9% versus SLE range 0.03-53.6%, median 9,75%, *p< 0.0001*) (**Fig. 1d**). Notably, SLE patients split almost evenly into patients with a low B_GC_ frequency comparable to HD (B ^LOW^), and ∼52% of SLE patients having a significantly higher numbers of B_GC_ (B ^HIGH^). SLE B_GC_ was also characterized by higher frequencies of Ki-67^+/hi^ B_GC_ Dark-Zone (DZ) cells (**Fig. 1e**). Isotype-characterization of the B_GC_ by flow cytometry revealed a predominant class-switching into IgG and a minor proportion of IgA-bearing cells in LNs, with the latter being slightly reduced in SLE (**Fig. 1f**). Concomitant evaluation of T helper cell subsets in LNs (**Fig.1 g-l**) also showed a significant increase of CXCR5^+/hi^PD-1^+/hi^ GC-T_FH_ restricted to SLE LN samples (*p = 0.001*) (**Fig. 1i**), and an even greater expansion of extra-follicular helper cells (T_EFH_, *p < 0.0001*), a population recently linked to GC-T_FH_^17^ and possibly Tph in blood^18^ (**Fig. 1l**). B_GC_ positively correlated with GC-T_FH_ in both HD and SLE (**Extended Data Fig. 1e**), but only SLE displayed a significant positive correlation between B_GC_ and T_EFH_ (**Extended Data Fig. 1e**). Consistent with LN structure changes with aging^19, 20^, we observed a negative correlation between B_GC_ frequencies and donor age (**Extended Data Fig. 1f**), a correlation which was clearer among SLE patients. However, SLE B_GC_ frequencies remained elevated relative to HD across the age spectrum.

### SLE lymph nodes accumulate heterogeneous plasma cell subsets enriched in mature ASC

We next addressed the composition of plasma cells (B_PC_) residing in human LNs. We first characterized early B_PC_ retaining the expression of CD19 (CD19^+^IgD^-^CD20^lo/-^CD38^hi^ cells). These cells expressed high levels of IRF-4 – a master transcription factor for B_PC_ – and low/mid-levels of Ki-67 (**Fig. 1k**). As observed for B_GC_, B_PC_ were greatly expanded in SLE LNs, while “resting” HD LNs had few to no B_PC_ (HD range 0.11-3.76%, median 0.87% versus SLE range 0.16-69.1%, median 5.8%, *p < 0.0001*) (**Fig. 1l**). SLE B_PC_ displayed significant heterogeneity as they included newly minted, Ki-67^+^ proliferative cells as well as mature PC expressing CD138 B_PC_ appeared to be composed by heterogenous cells that expressed both mature markers such as CD138^+^ B_PC_ (**Fig. 1m, n)**. LN B_PC_ retained some surface Ig, with IgG-expressing B_PC_ cells more frequent in SLE (**Fig. 1o**). Finally, to account for cells that retain some level of CD20, we assessed the overall frequency of CD27^hi^CD38^hi^ antibody-secreting cells (ASC) out of both CD19^+^ and also the more mature compartment of CD19^-^ cells (**Fig. 1p**). As previously observed for CD19^+^ B_PC_, all ASC from both CD19^+^ and CD19^-^ compartments were more frequently detected in SLE LNs (**Fig. 1q, r)**. These results suggest that lupus LNs are a site of newly-minted ASC generation as well as a niche for more mature plasma cells.

### SLE B_GC_ and B_PC_ are characterized by large clonal expansions with skewed isotype usage

We investigated the clonal composition of the human LN B cells using the high-throughput single-cell 10X platform to obtain ∼30,000 paired VH/VL sequences from two combined, independent single cell experiments. For the purpose of BCR clonal analysis, VDJ sequences were assigned to UMAP cell clusters defined by single-cell transcriptomics, which identified B_GC_ DZ and LZ, B_PC_ and non-B_GC_ cells separated into Naïve, CD27^+^ memory and CD27^-^ cells (**Extended Data Fig. 3a**). As shown in **Fig. 2a** by visualization of clonotype grouping^21^ of two representative honeycombs of HD-018 and SLE-022 subjects, SLE samples contained a higher number of expanded clones (**Fig. 2a, b** and **Extended Data Fig. 3b).** Clonal expansions were dominated by IgG1 isotype in SLE, with equal representation of IgM and IgG1 in HD. In addition, IgG4 was overrepresented in lupus (**Fig. 2c**). Large clones (n>11) were uniquely present in B_GC_ and B_PC_ in SLE, with some clonal families reaching above 50 members (**Fig. 2d**). Irrespective of clonal size, B_GC_ and B_PC_ had a greater representation of IgG1 then other compartments, including class-switched CD27^+^ Bmem (**Fig. 2e**). Moreover, the frequency of IgG1 was increased also in B_PC_ when compared to B_GC_ (**Fig. 2e**). As observed for clonal expansion, IgG4 was observed more frequently in B_GC_ and B_PC_ compartments (**Fig. 2e** and **Extended Data Fig. 3c**). When the overall diversity of the BCR repertoire was considered, the majority of SLE donors analyzed (62.5%), had a lower Shannon index and a higher Gini coefficient than HD, consistent with higher degrees of clonality and lower diversity (**Fig. 2f**). Notably, SLE subjects with oligoclonal repertoire belonged in the B_GC_^HIGH^ group. In HD, the majority of B_PC_ were clonally connected with IgG^+^ CD27^+^ memory cells (**Fig. 2g, h**). Conversely, in SLE the majority of B_PC_ were clonally connected with B_GC_ and/or B_Naive_. Consistent with GC-independent generation, approximately ∼20% of lupus B_PC_ were exclusively connected to B_Naive_ (**Fig. 2h**). Overall, these data demonstrate substantial differences in lymph node–resident B cell receptor architecture and selection between SLE and HD, and identify extensive clonal expansion and sharing between B_GC_ and B_PC_ compartments in lupus.

**Fig. 2.**
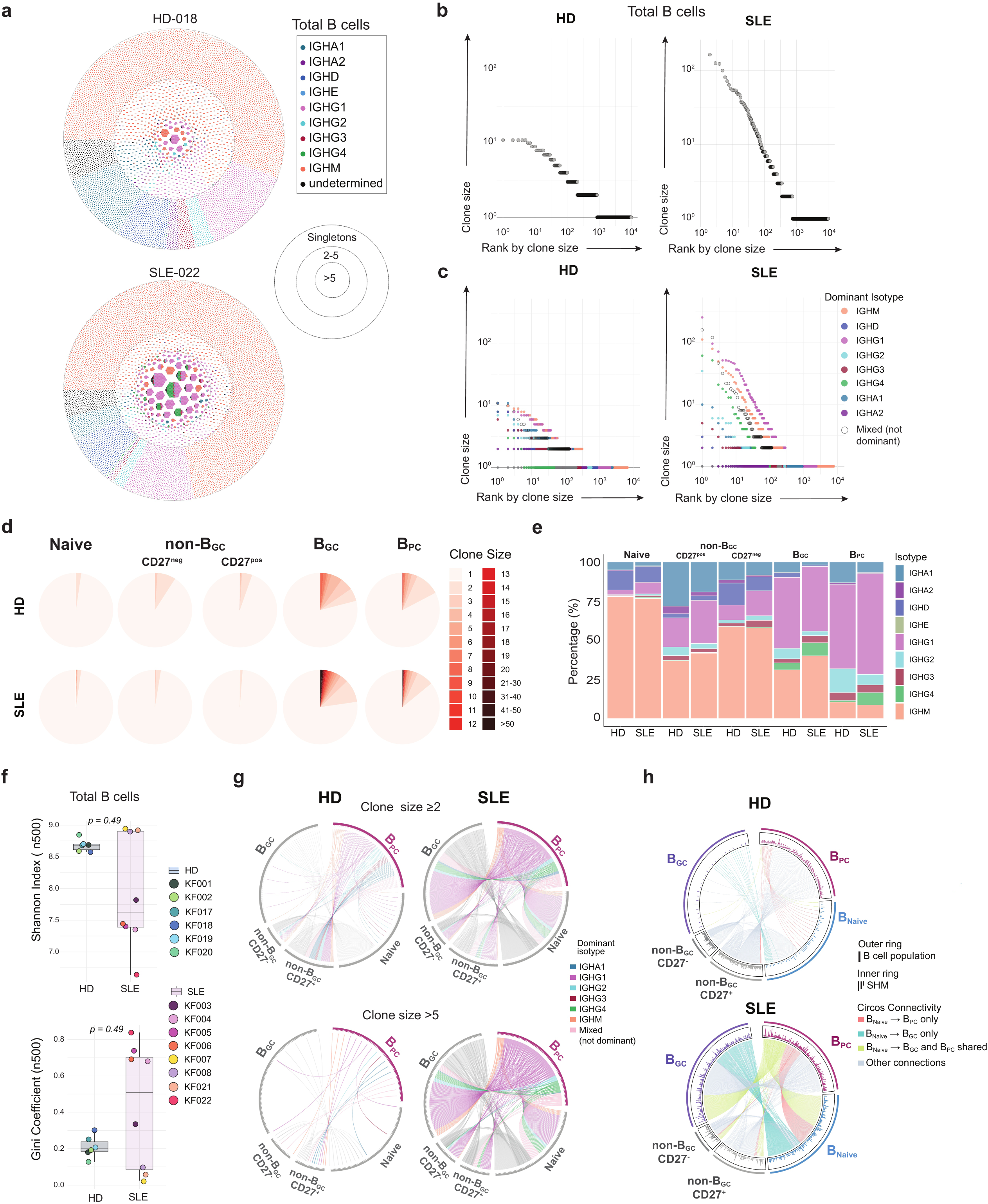
BCR structure of autoimmune SLE lymph nodes. **a,** Representative Honeycombs showing HD and SLE B cell clonal distribution (separated by singletons, small clones size 2-5, and large clones >5) and their isotype usage. **b,** Waterfall plots showing the clonal size and their ranking by size in HD and SLE. **c**, Waterfall plots showing the clonal size and their ranking by dominant isotype usage (at least 75% of isotype usage is considered dominant) in HD and SLE. Isotypes that did not reach at least 75% of dominance are shown as “Mixed” in the figure. **d**, Pie charts representing B cell clonal size distributions as identified in the indicated populations. **e**, Isotypes distribution used by the indicated B cell populations in HD and SLE. **f**, Shannon and Gini diversity indices calculated by random subsampling of n=500 clones in total LN B cells. Mann Whitney U test is shown for comparisons. **g,** Circos plots showing the clonal connectivity of B cell populations in HD and SLE, the top circos plots show the “clone size >=2” connections, the bottom plots show all “clone size >5” connections. The colors indicate each dominant isotype (75% or more) or the mixed ones (not dominant) and all the connections from Bnaive, non-B_GC_ CD27^-^, non-B_GC_ CD27^+^ and B_GC_ to B_PC_. **h**, Circos plots highlighting the connections of Bnaive to B_GC_ or B_PC_ exclusively, the shared between and the other connections for all VH repertoire.

### Autoreactive 9G4 and Ro60 B cells express high levels of PD-1 and Tox and progress uncensored in SLE B_GC_ and B_PC_ compartments

Next, we analyzed autoreactive B cells in the IgD^+^ compartment (CD21^+^CD11c^-^ resting Naïve “rB_N_” and CD21^-^CD11c^+^ activated Naïve “actB_N_”) as well as the IgD^-^ compartment. The latter was separated into B_PC_, B_GC,_ and non-B_GC_ B_CD27+_ memory and B_CD27-_ “double-negative” B_DN_ subsets (CD21^+^CD11c^-^ B_DN1,_ CD21^-^CD11c^+^ B_DN2,_ CD21^-^CD11c^-^ B_DN3,_ and CD21^+^CD11c^+^ B_DN4_) (**Extended Data Fig. 4a, b**).

The frequency of LN 9G4^+^ B cells (**Fig. 3a**), was determined in SLE donors that lacked 9G4 IgG seroreactivity, indicating preservation of tolerance (tolerant group, here referred to as SLE Seronegative “SeroNeg”), as well as in SLE patients with elevated 9G4 IgG seroreactivity (non-tolerant group, here referred to as SLE Seropositive “SeroPos”). We observed an increased proportion of 9G4^+^ cells in the actB_N_ of SLE donors, with higher frequencies in the “SeroPos” donors (**Fig. 3b**). SLE “SeroNeg” also showed an increase over HD. Consistent with the enforcement of downstream peripheral tolerance checkpoints, the “SeroNeg” patients did not expand 9G4^+^ B cells in GC and PC compartments. In contrast, “SeroPos” SLE had significant expansion of 9G4^+^ B cells in B_GC_ (**Fig. 3c)** and B_PC_ (**Fig. 3d**), alongside non-B_GC_ (**Fig. 3e).** Within the B_DN_ compartments (**Fig. 3f**)., SLE patients expanded 9G4^+^ B_DN2_ and B_DN3_. 9G4^+^ B_DN2_ were significantly more frequent in the SLE “SeroPos” group (mean HD ∼2% versus SLE “SeroNeg” ∼6.2% *pANOVA= 0.08*, HD versus SLE “SeroPos” ∼11.3% *pANOVA< 0.0001*, SLE “SeroNeg” versus SLE “SeroPos” *pANOVA= 0.03*), while 9G4^+^ B_DN3_ were increased in all SLE compared to HD (pANOVA <0.0001) (**Fig. 3f**). Similarly to 9G4 responses, Ro60^+^ B cells (**Extended Data Fig. 4c** and **Fig. 3g**) were significantly expanded in in the actB_N_ (**Fig. 3h**), as well as B_GC_ (**Fig. 3i**) and B_PC_ (**Fig. 3j**) in Seropositive donors. In this group, Ro60^+^ B cells were also increased in the B_DN_ non-B_GC_ (**Fig. 3k**), specifically with increased B_DN2_ and B_DN3_ cells in “SeroPos” SLE (**Fig. 3l**) (B_DN2_: mean HD ∼2% versus SLE “SeroNeg” ∼6.4% *pANOVA= 0.15*, HD versus the SLE “SeroPos” ∼13.12% *pANOVA< 0.0001*, SLE “SeroNeg” versus SLE “SeroPos” *pANOVA= 0.36*; B_DN3_: mean HD ∼8.4% versus SLE “SeroNeg” ∼22% *pANOVA= 0.12*, HD versus the SLE “SeroPos” ∼27.3% *pANOVA= 0.0014*, SLE “SeroNeg” versus SLE “SeroPos” *pANOVA=1*). Altogether, these results suggest that during autoreactive B cell engagement, tolerance failed in GC-mediated and PC-mediated checkpoints in SLE patients, with autoreactive B_PC_ expansion concomitant to an independent EF-signature (aN -> B_DN2/DN3_) prominent in Seropositive patients. Notably, when we analyzed the anti-Spike memory responses in our two cohorts, we did not observe differences in their distribution among various B cell compartments (**Extended Data Fig. 4d, e**). However, we still observed a higher frequency of non-B_DN1_ memory Spike in SLE patients (**Extended Data Fig. 4e**), as previously reported in PBMC upon mRNA vaccinations^22^, thus suggesting that the propensity to differentiate into non-B_DN1_ is a property of antigen-selected B cells associated with lupus-disease also in tissue. Strikingly, self-reactive 9G4- or Ro60-positive B cells in SLE were marked by an increased surface expression of PD-1 (**Fig. 3m, n**), and those PD-1^+/hi^ self-reactive cells also expressed higher levels of TOX (**Fig. 3o-p**). Those PD-1^+/hi^ TOX^+/hi^ B cells were not detected in anti-viral anti-spike memory B cell responses (**Fig. 3n, p**). Collectively, these results define unique properties of self-reactive B cells in SLE, capable of differentiating uncensored into germinal centers and of maturing as terminally differentiated plasma cells (**Fig. 3q**), selectively upregulating PD-1 and TOX and thus escaping checkpoint tolerance mechanisms that are instead in place in healthy donors’ LNs.

**Fig. 3.**
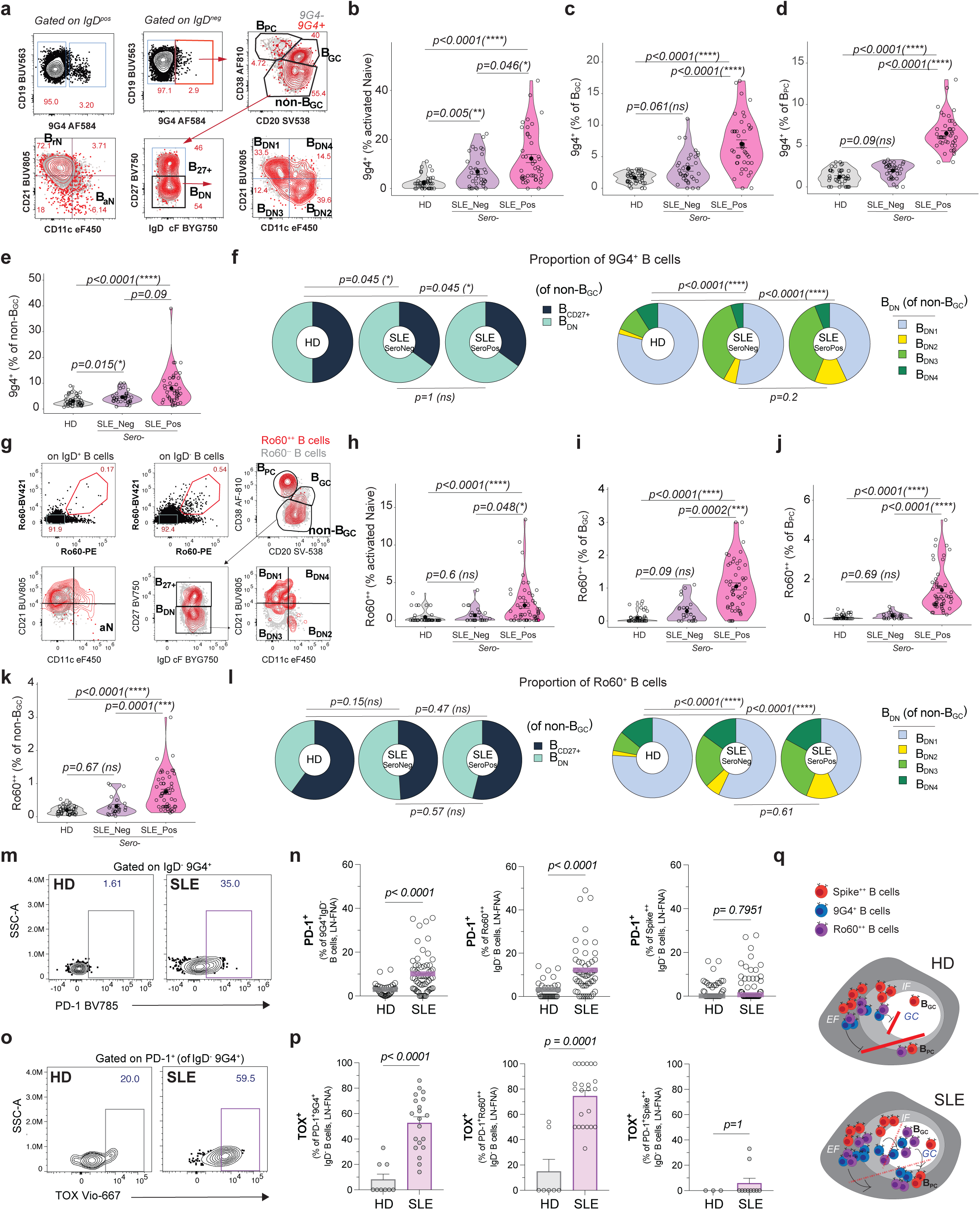
Tolerance escape of autoreactive B cells in SLE lymph nodes. **a,** Dot plots representative of the gating strategy to identify 9G4^+^ B cells among B cell populations in LNs. **b,** Violin plots representing the frequency of 9G4^+^ B cells gated on IgD^+^CD21^-^CD11c^+^ activated Naïve B cells. **c,** Violin plots representing the frequency of 9G4^+^ B cells gated on IgD^-^CD38^++^CD20^++^ B_GC_. **d,** Violin plots representing the frequency of 9G4^+^ B cells gated on IgD^-^CD38^++^CD20^-^ B_PC._ **e,** Violin plots representing the frequency of 9G4^+^ B cells gated on IgD^-^CD38^-^CD20^+^ non-B_GC_. Each dot represents an individual sample. Groups are shown as HD, SLE_Neg (9G4 IgG Seronegative) and SLE_Pos (9G4 IgG Seropositive). One-Way ANOVA test applied. **f,** Proportions of B_CD27+_ and B_DN_ gated on 9G4^+^ non-B_GC,_ and proportions of B_DN_ subsets (B_DN1-4_). Fisher’s exact test is shown among groups. **g,** Dot plots representative of the gating strategy to identify Ro60^++^ B cells among B cell populations in LNs. **h,** Violin plots representing the frequency of Ro60^++^ B cells gated on IgD^+^CD21^-^CD11c^+^ activated Naïve B cells. **i,** Violin plots representing the frequency of Ro60^++^ B cells gated on IgD^-^CD38^++^CD20^++^ B_GC_. **j,** Violin plots representing the frequency of Ro60^++^ B cells gated on IgD^-^CD38^++^CD20^-^ B_PC._ **k,** Violin plots representing the frequency of Ro60^++^ B cells gated on IgD^-^CD38^-^CD20^+^ non-B_GC_. Each dot represents an individual sample. Groups are shown as HD, SLE_Neg (Ro60 IgG Seronegative) and SLE_Pos (Ro60 IgG Seropositive). One-Way ANOVA test applied. **l,** Proportions of B_CD27+_ and B_DN_ gated on Ro60^++^ non-B_GC,_ and proportions of B_DN_ subsets (B_DN1-4_). Fisher’s exact test is shown among groups. **m,** Representative dot plots showing the expression of PD-1 on 9G4^+^ non-naïve B cells. **n,** Quantitation of PD-1 positive cells gated on 9G4^+^, Ro60^++^ or Spike^++^ non-naïve B cells in HD and SLE groups. Each dot represents an individual sample. Mann Whitney U test shown for comparisons. **o,** Representative dot plots showing the expression of intracellular TOX on PD-1^+^9G4^+^ non-naïve B cells. **p**, Quantitation of TOX positive cells gated in PD-1 positive 9G4^+^, Ro60^++^ or Spike^++^ non-naïve B cells in HD and SLE groups. Each dot represents an individual sample. Mann Whitney U test shown for comparisons. **q**, Simple cartoon model showing the “break-of-tolerance” to auto-antigens in SLE LNs and tolerance checkpoints in HD LNs.

### Loss of VH_4-34_ tolerance in SLE linked to expansion of low-mutated oligoclonal families that escape germinal center and plasma cell selection

Single-cell VDJ sequencing analysis allowed us to characterize *in depth* the properties of VH_4-34_-bearing B cells in our two cohorts. Mutations levels among VH families varies in SLE and according to B cell populations (**Extended Data Fig. 5a**), with certain families being more mutated (e.g., IGHV_1-69_, IGHV_4-4_) and other families characterized by lower mean mutation levels (e.g., IGHV_3-13_, IGHV_3-33_, IGHV_4-34_), relative to the same VH in the HD. To better understand the use of IGHV_4-34_ in our cohorts, when clonotype grouping of VH_4-34_ was restricted to the “Top10 usage” (**Extended Data Fig. 5b**), only one HD subject showed VH_4-34_ usage (n=1/6, 16.7%) contrary to SLE patients where half of the group showed VH_4-34_ in the Top10 list (n=4/8, 50%). Importantly, the utilization of VH_4-34_ by LN B cells in SLE correlated with 9G4 seropositivity (**Extended Data Fig. 5c**). Increasing the ranking to “Top20 usage” (**Fig. 4a**) expanded the detection of VH_4-34_ also in the HD group, with n=5/6 subjects ranking its usage from positions 11^th^ to 15^th^, and n=1/6 ranking 3^rd^. Similarly, 50% of SLE subjects (n=4/8) ranked above the 10^th^ position (from 4^th^ to 8^th^), and 50% (4/8) ranked from 13^th^ to 18^th^ position in the Top20 analysis (**Extended Data Fig. 5d)**. Representative honeycombs plots are shown in **Fig. 4a**. Both the number of total VH_4-34_ clones as well as the expanded ones (n= ≥2 cells) (**Fig. 4b**) were slightly higher in SLE (n=468 and n=596 total clones; n= 31 and n=36 expanded clones in HD and SLE, respectively), with higher total clone counts in SLE B_Naïve_, and overall clonal enrichment in SLE B_GC_ (n= 11 and n= 63 total clones, n= 3 and n= 23 expanded clones in HD and SLE, respectively) and B_PC_ (n= 8 and n= 56 total clones; n= 1 and n= 18 expanded clones in HD and SLE, respectively) (**Fig. 4b**). Large VH_4-34_ clones (size >10) were mostly present in SLE B_GC_ and B_PC_ (**Fig. 4c**). Isotype usage of VH_4-34_ clones largely reflected the overall distribution of Ig previously observed in the full VH repertoire (**Fig. 4d)**. Overall, analysis of SHM by B cell subsets showed reduced mutation levels in lupus VH_4-34_ B cells, particularly for those expanded clones residing in B_GC_ and B_PC_ (**Fig. 4e**). While in HD the VH_4-34_ class had little to no clonal sharing, SLE patients had more inter-clonal connections, with their B_Naive_ contributing to exclusive and shared B_GC_ and B_PC_ clonal connections (**Fig. 4f)**. Unsupervised metadata clustering of samples based their 9G4 seroreactivity associated their blood and LNs immune cell populations and VH_4-34_ properties and separated the subjects into two distinct clusters. The 9G4 Seropositive SLE associated with activated immune cells – including both blood and LN actB_N_, B_DN2/3_, B_GC,_ and B_PC_, as well as LN GC-T_FH_ and PD-1^+^CXCR5^-^ T_EFH_ helper cells – alongside an overall reduced level of VH_4-34_ SHMs (**Fig. 4g**). Conversely, SLE 9G4-SeroNegative donors clustered with HD and showed instead higher VH_4-34_ mutation levels (**Fig. 4g**). Altogether, these data provided original information on VH_4-34_ B cells distribution in LN B cells, their selection and expand beyond 9G4 positivity their lack of censoring mechanisms in the GC (**Fig. 4h**).

**Fig. 4.**
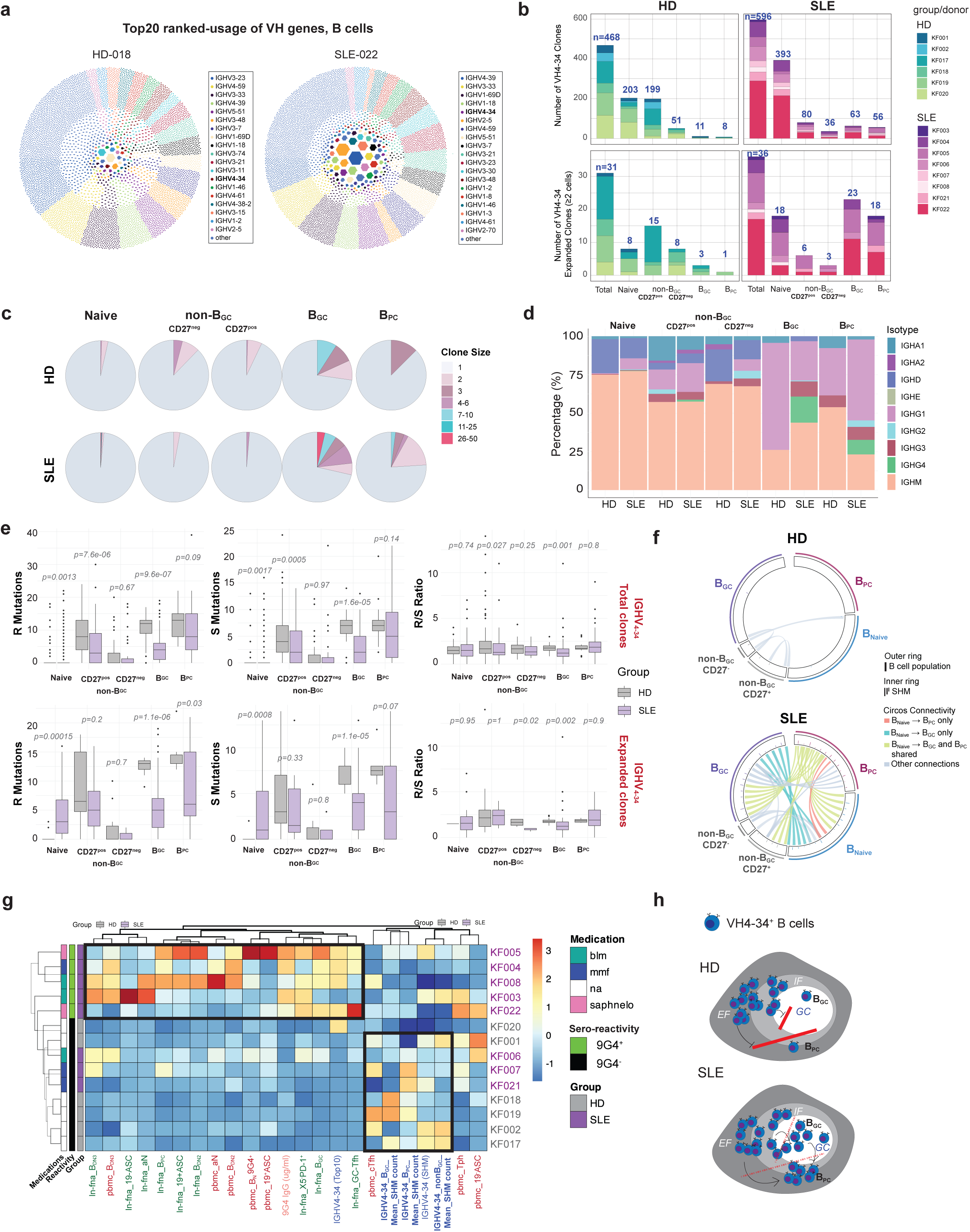
VH_4-34_-bearing B cells characterization in human lymph nodes. **a,** Representative Honeycombs showing HD and SLE B cell clonal distribution (separated by singletons, small clones size 2-6, and large clones >6) and their top-20 ranked-usage of VH genes. **b,** Quantitation of total VH_4-34_ clones (top bars) and expanded clones (two or more cells) in all LN B cells or the indicated B cell populations. The numbers indicate the number of reads. Each donor is indicated by different colors. **c**, Pie charts representing the clonal size of VH_4-34_ in the indicated B cell populations. **d**, Isotypes distribution used by the indicated B cell populations in HD and SLE. **e**, SHM count in both total and clonally expanded VH_4-34_ in the indicated populations. Replacement (R), Silent (S) and R/S ratio are shown. Mann Whitney U test shown for comparisons. **f**, Circos plots highlighting the connections of Bnaive to B_GC_ or B_PC_ exclusively, the shared between and the other connections for VH**_4-34_** repertoire. **g**, Heatmaps showing the HD and SLE subjects tested by single cells and separated by 9G4 seropositivity. Cluster analysis shows the association with immune signature in blood and lymph nodes, as well as levels of SHM in the VH_4-34_ gene. **h**, Simple cartoon model showing the “break-of-tolerance” to VH_4-34_ in SLE LNs and tolerance checkpoints in HD LNs.

### “Clonal damnation” of VH_4-34_ clones in SLE allows self- and polyreactive B cells to escape tolerance, acquire mutations, and proceed unselected “towards-self”

To gain insights into the mechanisms regulating B cell tolerance in SLE, we first analyzed the mutations occurring in the VH_4-34_ BCRs, and particularly in the A^23^V^24^Y^25^ region, as part of the FR1 unmutated hydrophobic patch “HP” that retains self-reactivity in 9G4^+^ B cells^9^ (**Fig. 5a**). LN B cells from SLE donors showed lower levels of mutations of the “QW--AVY---NHS” regions (**Fig. 5a, b**), with progressive loss of germline-like properties when following B cell differentiation from Naïve to mature subsets (**Fig. 5b**). Notably, while HD VH_4-34_ AVY^+^-BCRs underwent extensive SHM when expressed on B_PC_ (∼77%), SLE VH_4-34_ AVY mutations were unchanged in both B_GC_ (∼44%) and B_PC_ (∼42%) compartments (**Fig. 5a, b**), suggesting that those SLE VH_4-34_ AVY^+^-BCRs were not subjected to “clonal redemption” by SHM as observed in healthy donors^13^. One representative possible escape from “clonal redemption” in SLE of the “AVY patch” was identified in the clonal family “6783” isolated from subject “SLE-004”, allocating large size members (clonal size n=30) and in which all clonal members maintained low levels of mutations and all-but-one retained the AVY patch, while differentiating in several B cell compartments (**Fig. 5c**). To better define the binding properties of tissue-resident VH_4-34_-expressing B cells, we selected VH_4-34_ clones from both cohorts and expressed them as monoclonal antibodies (mAbs). These mAbs were tested against a broad panel of self-antigens (HEP-2, dsDNA, Ro60, Ro52, Smith, H3, La, LPS and PCNA) and viral antigens (EBNA1 and Wuhan.1 Spike trimer). We additionally performed flow assays to assess their cellular binding to HD naïve B cells (B-cell binding, BCB; **Extended Data Fig. 6a**). Results were compared with the strongly polyreactive VH_4-34_ mAb 75G12, previously isolated from circulating CD27⁺ memory SLE B cells^9^. A representative example for LN-derived VH_4-34_ mAbs (**Fig. 5d**), shows clones exhibiting BCB activity (mAbs #4 and #5) as well as a clone lacking this property (mAb #8). 9G4^+^ mAb have been reported to specifically bind B cells via anti-i B cell binding (BCB)^9^, but they also share other autoreactivities with the 9G4^-^ AVY-mutated mAb such as binding to anti-nuclear antigens or apoptotic cells^9, 23^. Thus, we used J45.01 *CD45-knock-out* Jurkat T cell line pre-treated with Camptothecin, in order to induce apoptosis (Apoptotic-cell binding “APCB”, **Extended Data Fig. 6b**). Interestingly, B_GC_ are formed by highly proliferating and cycling B cells that undergo high-rates of B-cell apoptosis^24, 25^ and human SLE B_GC_ showed a higher frequency of active-caspase3/7-positive cells (**Fig. 5e**), a possible source of self-derived apoptotic cell components. Similar to BCB, also APCB was detectable in some VH_4-34_ clones, such as 75G12, mAbs #4 and #5 (**Fig. 5f**). Notably, mAbs #4 and #5 belonged to the clonal family “7120” isolated from subject “SLE-005” (**Fig. 5g**) and represented medium-size clones (clonal size n=9) that retains self-reactivity despite accumulating mutations in both B_GC_ and B_PC_. Both clones from “6783” and “7120” families retained various degrees of self-reactivities when tested for their binding to Hep-2 cells (**Fig. 5h**), and recapitulated similar staining patterns to the sera reactivity observed in the donors from which they were isolated (**Fig. 5h**). Additionally, since mAbs #4 and #5 differed from a few mutations in the V_HC_, but carried also different V_LCs_, and to further investigate the contribution of V_LCs_ to autoreactivity, we performed LC-swapping experiments (**Fig. 5i**). Overall, we observed that intra-clonal swapping (e.g., 4HC-5LC, 5HC-4LC, 6HC-4LC)) mostly maintained the original self-reactive properties, with minor modifications to reactivity intensity or binding. Inter-clonal swapping with non-self-reactive V_LCs_ (e.g., 4HC-8LC, 5HC-8LC) still retained some self-reactivity, while the swapping with self-reactive V_LCs_ (e.g., 8HC-4LC, 8HC-5LC or 13HC-5LC) only minimally increased the self-reactivity, with both cases failing to carry the full self-reactive properties of the strongly self-reactive mAbs. Overall, these results support evidence for which the right combination of V_HC_ and V_LC_ is necessary to maintain high levels of self- and poly-reactivity, but the V_HC_ contributes largely to the strong autoreactive properties of the VH_4-34_ BCR (**Fig. 5i**).

**Fig. 5.**
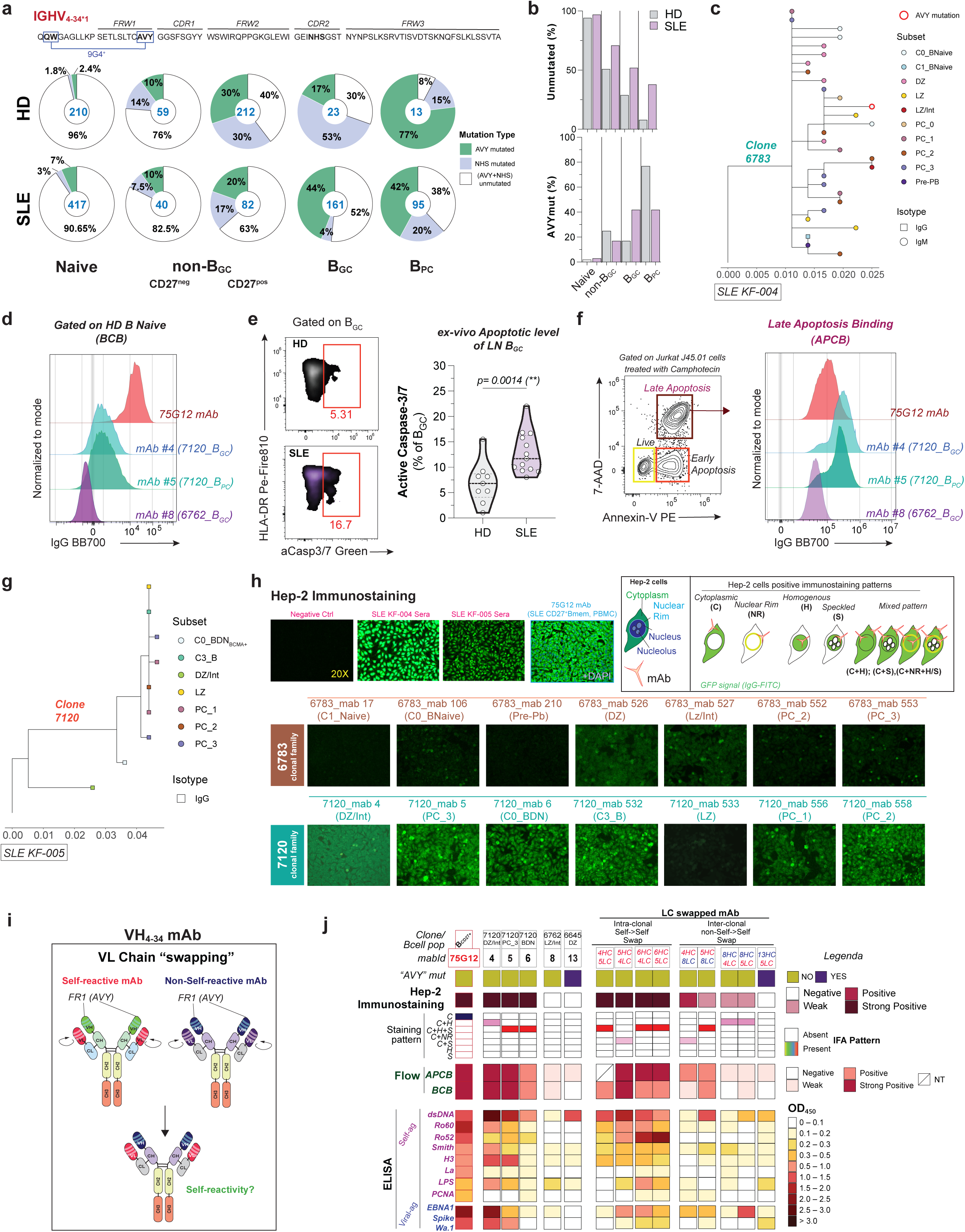
“Clonal damnation” of SLE VH_4-34_ clones. **a,** Sequence of VH_4-34_ gene and pie charts showing the frequency of unmutated AVY+NHS, NHS-only mutated or AVY-only mutated regions in VH_4-34_ sequences from the indicated B cell populations in HD and SLE. The numbers in the pies represent the number of sequences detected by 10X VDJ repertoire single cell analysis. **b,** Bar plots showing the percentages of unmutated (top bars) and AVY-mutated (bottom bars) sequences in HD and SLE. **c**, Phylogenetic tree representing clonal family 6783 showing members isolated from donor SLE-004. B cells subsets are indicated by color, isotype by shape. Avy mutation in a red circle. Mutation frequencies are shown in the x-axis. **d**, Representative dot plots for BCB flow assay showing mAb binding detected with IgG positivity on healthy donor Naïve B cells. IgG range: Negative values 0-3%, Weak values 4-8%, Positive values 9-30%, Strong positive values >30%. **e**, Representative dot plots showing the percentage of apoptotic cells as Caspase3/7-positive gated on B_GC_ cells and their quantitation in violin plots. Each dot represents an individual sample. Mann Whitney U test shown for comparison. **f**, Representative dot plots for APCB flow assay showing mAb binding detected with IgG positivity on Annexin V and 7-AAD positive Jurkat cells. IgG range: Negative values 0-5%, Weak values 6-10%, Positive values 11-30%, Strong positive values >30%. **g**, Phylogenetic tree representing clonal family 7120 showing members isolated from donor SLE-005. B cells subsets are indicated by color, isotype by shape. Mutation frequencies are shown in the x-axis. **h**, Hep-2 immunostaining of the indicated SLE sera or mAbs from the two clonal families shown in the trees (**c, g**). The cartoon represents the common staining patterns identified upon incubation of a mAb with Hep-2 cells and its detection with anti-IgG FITC. **i**, Carton representing the VL light chain swapping by transfecting different variable chains into selected mAbs to assess the changes in self-reactivity binding properties **j**, Heatmap showing the identification of each indicated mAb (*mabID*), its clone id and B cell population (Clone/B cell pop), and if AVY-patch was germline, unmutated (NO) or mutated (YES). Values and ranges are indicated in the legenda. Help-2 immunostaining and patterns, flow of BCB and APCB assays, and ELISA for self- (dsDNA, Ro60, Ro52, Smith, H3, La, LPS, PCNA) or viral- (EBNA1, Spike) are shown.

### VH_4-34_ antibodies from SLE exhibit broader self- and viral-antigen cross-recognition

Screening of autoreactivity of a large panel of VH_4-34_ mAbs isolated from both HD (n=41 mAbs) and SLE (n=81 mAbs) allowed us to compare their binding and molecular properties. Although the frequency of VH_4-34_ clones in germinal center and plasma cell compartments differed of a six-fold factor between HD and SLE (**Fig. 4b**), the overall analysis of Hep-2 reactivity for the isolated mAbs showed similar reactivity levels between the two groups (**Fig. 6a**). However, the IFA staining pattern was able to distinguish the groups, with HD mAbs accumulating more cytoplasmic staining, while SLE mAbs characterized by increased homogenous and mixed patterns (**Fig. 6a**). Germline AVY-patches versus the mutated ones did not differ in HD, but associated with more Hep-2 reactivity in SLE (**Fig. 6a**), similar to stronger BCB and APCB overall observed in the AVY-unmutated mAbs (**Fig. 6b**). At the molecular level, SLE mAbs had distinct features, with reduced level of SHM and despite increased self-reactivity (**Extended Data Fig. 6c** and **Fig. 6c**), and overall distinct properties related to the mutational sites as well as CDR3H properties in the groups (**Extended Data Fig. 6d** and **Fig. 6d**). Notably, strongly positive mAbs in SLE carried longer and more mutated CDR3Hs. These strongly positive mAbs also exhibited the highest *TOX* transcript levels (**Fig. 6e**), with *TOX* expression progressively increasing with the strength of Hep-2 binding (**Fig. 6e**). The higher *TOX* transcript levels in self-reactive mAbs were consistent with the elevated TOX protein expression observed on PD-1⁺ 9G4 B cells in SLE reported in Figures 4m and 4n. Broad anti-self and anti-viral reactivity was assessed for some of those mAbs (**Extended Data Fig. 7** and **Fig. 6f**), confirming previously reported binding of VH_4-34_ to nuclear antigens such as dsDNA^9^ or LPS^26^, but also expanding their binding properties to anti-viral proteins such as the Spike or the structural EBV protein EBNA1(**Fig. 6f**). Overall, most of those strong self-binders retained the germline-AVY patch and showed reduced SHM levels (**Fig. 6f**), as previously observed (**Fig. 6a-c**). Exclusively in SLE, a group of mAbs exhibit strong poly-reactive properties (**Fig. 6f-h**). Of those strong self-reactive mAbs in SLE, ∼8% of GC- or PC-derived mAbs carried cross/poly-reactivities that included both anti-self and anti-viral antigens (**Fig. 6h**).

**Fig. 6.**
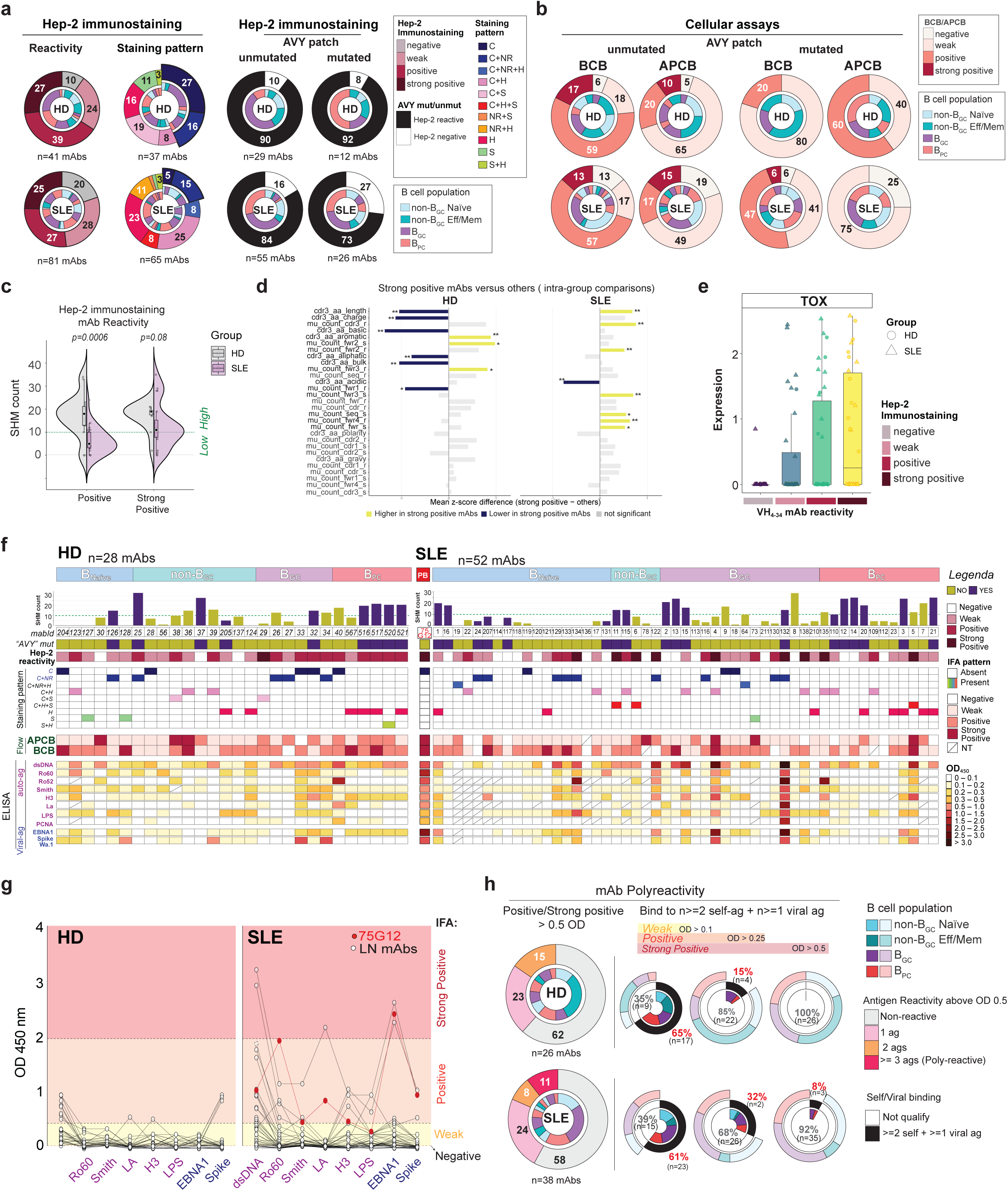
Increased polyreactivity and distinct selection of SLE VH_4-34_ mAbs. **a,** Pies showing the combined results from Hep-2 immunostaining for: type of reactivity, staining patterns and frequency of self-reactivity in AVY-unmutated and AVY-mutated mAbs. **b**, Pies showing the combined results for cellular assay reactivity distribution. BCB and APCB separated by AVY mutation state. Inner donuts show the respective B cell populations as indicated in the legenda. **c,** Scatter box violin plots showing the SHM count (R+S) levels in the mAbs from HD or SLE that bound to Hep-2 with positive and strong positive signals. Mann-Whitney U Wilcoxon rank-sum test between HD ad SLE group is shown. Boxes indicate the medians. **d**, Diverging bar plots showing HD and SLE tested mAbs in Hep-2 immunostaining and their features. Mean values are z-score transformed prior to plotting. Kruskal-Wallis’s statistics indicated when significant among the four IFA Hep-2 immunostaining groups (negative, weak, positive, and strong positive). **e**, TOX transcripts expression on mAbs from HD and SLE separated based on Hep-2 reactivity. **f**, Heatmap showing the identification of each indicated mAb (*mabID*), SHM counts, and if the AVY-patch was germline, unmutated (NO) or if it was mutated (YES). Values and ranges are indicated in the legenda. Hep-2 immunostaining and patterns, flow of BCB and APCB assays, and ELISA for self-antigens (dsDNA, Ro60, Ro52, Smith, H3, La, LPS, PCNA) or viral-antigens (EBNA1, Spike) are shown. **g**, Graphs showing mAbs from HD and SLE tested in ELISA for all 8 antigens. Each dot represents an individual mAb, and connecting lines shown in the same mAb tested for multiple antigens. Arbitrary ranges based on ELISA O.D. values are shown in the graph. **h**, Analysis of polyreactivity (here refer to as the binding to three or more antigens). Donuts showing the level of poly-reactivity from mAb tested for all eight antigens by ELISA. HD (n=26) and SLE (n=38) are compared for signals above O.D. 5 (positive and strong signals) and for cross-poly-reactivity to at least two self-antigens and one viral antigen, shown for OD values above 1, 2.5 and 5. Inner donuts show the respective B cell populations as indicated in the legenda.

## Discussion

In this study, we sought to characterize lymph node–resident B cells in autoimmune lupus subjects and healthy controls and interrogate their reactivity against self-antigens relevant to lupus disease. Cellular therapies have recently shown remarkable efficacy in SLE, suggesting that deep B cell depletion and immune resetting may be critical for achieving long-term remission and drug-free states. Analyses of secondary lymphoid organs after CAR-T therapy have reported follicular disruption and near-complete B cell depletion^27^, underscoring the pathogenic relevance of lymphocytes primed within lymph nodes in lupus. Notably, a recent *proof-of-concept* study proposed the use of VH_4-34_ targeted CAR-T (CAR4-34)^28^ to specifically target self-reactive B cells in lupus and cancer, reducing the risk of antigen-negative escape observed at times with CAR-19^29^. Given the impact of these findings, it is imperative to address gaps in our understanding of the cellular composition and immune regulation of tissue-resident B cells in human LNs, thereby establishing a baseline reference for comparison with therapeutic interventions.

Access to human LNs from SLE patients therefore provides a unique opportunity to profile the overall immune landscape of lymphoid tissues and study B cell receptors structure, as immune receptor repertoires can serve as indicators of an individual’s immunological state^30^. Additionally, it allows us to interrogate specifically the compartment of antigen-specific B cells and directly examine how self-reactive clones are distributed across B cell subsets compared with non–self-reactive populations.

Here, despite enrolling SLE subjects with low disease activity, we observed spontaneous activation of LN B_GCs_ with higher levels of apoptosis^24^, and a concomitant expansion of PD-1–positive T cell subsets (T_EFH_ and GC-T_FH_), suggesting that, in the absence of infections or vaccinations and with controlled clinical activity, secondary lymphoid tissues in lupus are chronically exposed to antigen-driven immune responses, likely through continuous exposure to self-antigens. Alongside hyperactive germinal centers, SLE LNs hosted a greater number of terminally differentiated plasma cells. These data are consistent with the reported expansion of blood-circulating B_PC_ in SLE with a more mature phenotype^2, 3^, and might suggest that LNs could represent a niche for the accumulation of tissue-resident plasma cells with pathogenic potential and contribution to tissue damage.

We observed a larger oligoclonal expansion in SLE LN B cells, with altered isotype usage and a greater representation of IgG4-bearing BCRs. While pathogenic IgG4 autoantibodies are well reported in various autoimmune conditions, such as IgG4-RD, Pemphigus, and Myasthenia gravis^31, 32^. IgG4 B cells are not usually detected in studies with peripheral blood. We now showed that IgG4 B cells are detectable with significant frequencies in lupus LNs and associated with autoreactivity.

This study addressed the regulatory peripheral B cell checkpoints existing in humans and healthy states, and how in lupus those mechanisms fail to remove autoreactivity. Overall, our study revealed how two unrelated lupus antigens escaped B cell tolerance and progressed unimpeded in the GC and PC compartments. Interestingly, SLE donors seropositive for the two self-antigens accumulate also increased numbers of antigen-reactive CD21^lo^CD11c^+^ activated naïve, a finding consistent with epigenetic programs underpinning B cell dysfunction in SLE at the naïve stage^33^. Additionally, these donors also presented expanded self-reactive B_DN2_ and B_DN3_ cells, and B_PC_ originating from naïve cells, suggesting that alongside persistent GC activity, breach of B cell tolerance to self-antigens might co-exist with *de novo* extrafollicular autoreactive plasma cells formation^34, 35^. When compared to anti-viral memory responses, we observed a clear distinction with 9G4 and Ro60 self-reactive B cells in SLE that expressed higher levels of PD-1 and TOX. Increased PD-1 expression has been reported both in lupus T^18, 36^ and B cells^37^, with PD-1 expression linked to self-reactive T cells^38^. While the exact mechanism of regulation of those PD-1^+/hi^Tox^+/hi^ B cells in SLE still remains to be elucidated, we propose that chronic autoantigen exposure and sustained BCR signaling might be responsible for this mechanism and thus distinguish the selection of self-reactive B cells in SLE when compared to both HD-reactive B cells as well as anti-viral B cells. We further assessed *in depth* the phenotype and properties of VH_4-34_ B cells bearing the 9G4 idiotype (9G4^+^ B cells), which has been widely demonstrated to represent a useful tool to study tolerance regulation in lupus^9^. Additionally, this study provides a parallel comparison with VH_4-34_ B cells that have lost the hydrophobic AVY patch in the FW1 region of their V_HC_ and therefore cannot be identified by 9G4 probing (9G4^−^ B cells). VH_4-34_ belongs to a VH gene family that shows low levels of polymorphism across the general population, suggesting that this gene is under strong selective pressure to be maintained and participate in immune-mediated responses. In healthy conditions, VH_4-34_ antibodies are regulated by process of clonal redemption^13^ and might be important for their participation to protective immune responses, such as anti-microbial or anti-viral responses, as reported by their broad recognition of commensal bacteria^39^ or vaccinia virus responses^13^. In SLE, 9G4^+^ B cells persisted and expanded as unmutated clones, and represent a significant fraction of anti-native double-stranded DNA (anti-ds-DNA) antibodies– carrying a disease specificity of ∼95% and correlating with disease severity – while also representing the majority of anti-B cell CD45 antibodies, suggesting that the mechanism of self-reactive clonal redemption might be at fault in autoimmune settings, and instead progress unimpeded into maturation and generation of autoreactive plasma cells with pathogenic potentials. We provided original evidence showing that B cell selection and aberrant affinity maturation processes are responsible for self-reactive clonal escape to tolerance checkpoint in the B_GC_, with consequent accumulation of mutations in the B_PC_. This mechanism, hereby referred to as “clonal damnation”, proposes a system where normal selection of self-reactive clones of the VH_4-34_ family against self or “clonal redemption”^13^ reported in healthy individuals, is faulty in SLE. Clonal members of families “6783” and “7120”, isolated from two different SLE patients, reflect this faulty mechanism where progressive maturation in B cell compartments, and accumulation of somatic hypermutations, favor the persistence of auto- and poly-reactive clones that escape tolerance checkpoints, and reside in GC and PC compartments, in which progressive accumulation of mutations failed to eliminate self-reactivity and instead preserved – or even reinforced – it. Noteworthy, somatic hypermutations appear to be critical in the VH_4-34_ selection process, with evidence of lower somatic mutations in SLE donors that had breached tolerance to 9G4 and carried polyreactive and clonally expanded VH_4-34_ clones in their LNs. While accumulating mutations could be detected in the same clones progressing towards maturation, the lower stringency of affinity maturation of the VH_4-34_ locus might suggest impaired affinity maturation and selection processes in lupus LNs, leading to expanded B_GC_ yet inefficient B cell selection. This would be consistent with both impaired tolerance to self-antigens as well as critically inefficient selection of protective immune responses as previously observed in vaccination settings^22^. Assessment of VH_4-34_ monoclonal antibodies binding against a broad panel of self- and viral-antigens revealed overall autoreactive properties, consistent with the VH_4-34_ BCR, extending to the previously reported properties of 9G4 idiotype B cells possessing autoreactivity against N-acetyl-lactosamine (NAL) determinants expressed by the l/i blood group antigens and other self-glycoproteins including CD45. As normal anti-glycan reactivity is linked to B cells that undergo affinity maturation in the germinal centers^40^, here we report a GC-mediated regulation of a BCR that retains anti-glycan reactivity but increases its polyreactivity towards other disease-specific autoantigens as well as viral antigens in pathogenic conditions. Indeed, SLE derived VH_4-34_ mAbs displayed higher levels of polyreactivity when tested against three or more self-antigens, and also cross-reactivity with at least one of our viral antigens (EBNA1 or Spike). These data support recent evidence in human viral infections where chronic exposure to SARS-CoV2 induced autoimmune-like symptoms that persisted even after viral clearance^41, 42^. Notably, one of the viral antigens strongly recognized by our SLE mAbs was EBNA1, a critical component of the Epstein-Barr virus (EBV) often reported in association with SLE^43, 44^. This study provides a mechanistic explanation that may underly this association.

## Supporting information

Ext Data

## Data Availability

All data produced in the present study are available upon reasonable request to the authors

## Extended Data figure legends

**Extended Data Fig. 1 Study cohort characteristics. a,** Scatter plots showing the cellular recovery of viable cells obtained with LN-FNA biopsy. Each dot represents an individual sample. Mann Whitney U test shown for comparisons. **b,** Cartoon showing the experimental design for the selection of de-identified donors analyzed with single cell 10X chromium assays. **c**, Table showing the characteristics of HD and SLE subjects selected for 10X single cell studies. Selected information for demographics, serology, and flow data from LNs and blood are shown. **d**, Box plot showing the available SLEDAI-SELENA score for the SLE donors enrolled in the study. Score 0-4 is considered low disease activity; score above 4 is considered high disease activity state. **e,** Graphics showing the correlation between B_GC_ and GC-T_FH_ (left), B_GC_ and T_EFH_ (middle), and GC-T_FH_ and T_EFH_ (right). **f,** Graphics showing the correlation between B_GC_ and age (years). Each graph includes the calculated Spearman correlations, shown for HD (grey line) and SLE (purple line) groups. Each dot represents an individual sample.

**Extended Data Fig. 2 Anti-self and anti-viral serological profiling of circulating antibodies. a,** Scatter plots showing the anti-Ro60 IgG levels quantitated with LIPS in HD and SLE sera. Data are represented as Arbitrary Units (A.U.), and horizontal line at 1×10^3^ represents the threshold for positive values. Each dot represents an individual sample. Mann Whitney U test shown for comparisons. Percentage in the plot indicates the frequency of SLE positive donors (∼72.2%). **b,** Scatter plots showing the anti-9G4 IgG levels quantitated with ELISA in HD and SLE sera. Data are represented as concentration (ug/ml), and horizontal line at 200 ug/ml represents the threshold for positive values. Each dot represents an individual sample. Mann Whitney U test shown for comparisons. Percentage in the plot indicates the frequency of SLE positive donors (∼53.2%). **c,** Scatter plots showing the Wuhan anti-Spike S1, S2 and RBD IgG levels quantitated with Luminex in HD and SLE sera. Data are represented as MFI values. Each dot represents an individual sample. Mann Whitney U test shown for comparisons. **d**, Scatter plots showing the Wuhan anti-Nucleocapsid IgG levels quantitated with Luminex in HD and SLE sera. Data are represented as MFI values. Each dot represents an individual sample. Mann Whitney U test shown for comparisons. **e,** Correlation between HD sera values for anti-S1 and anti-RBD IgG titers (MFI) and the same samples tested on SPR for affinity binding properties of polyclonal Ig to SARS-CoV2 wild-type S1 and RBD proteins. Each dot represents an individual sample. Pearson test applied. **f,** Correlation between SLE sera values for anti-S1 and anti-RBD IgG titers (MFI) and the same samples tested on SPR for affinity binding properties of polyclonal Ig to SARS-CoV2 wild-type S1 and RBD proteins. Each dot represents an individual sample. Pearson test applied. **g**, Correlation between SLE sera values for anti-Ro60 titers (A.U.) and the same samples tested on SPR for affinity binding properties of polyclonal Ig to Ro60 protein. Each dot represents an individual sample. Pearson test applied.

**Extended Data Fig. 3 BCR properties of LN B cells in HD and SLE. a**, UMAP showing the clustering of LN B cells into three compartments of non-B_GC_, B_GC_ and B_PC_. Main gene identity expression level is shown in the Umaps on the right. **b,** Pie charts showing the clonal expansion in total B cells from LN samples tested by single cells 10X runs and showed by donors for both HD (n=6) and SLE (n=8) cohorts. **c,** Box plot showing the SHM count for Naïve B cells in HD and SLE groups. Mann Whitney U test shown for comparisons. **c**, Bar plots showing the distribution of isotypes in each indicated B cell populations when clustered by singletons (clone size =1), small (clone size =2-5), and larger (clone size 6-9 and >=10).

**Extended Data Fig. 4 Cellular characterization of antigen-specific B cells in LNs. a,** Representative FACS plots showing the gating strategy used to identify the frequency of 9G4 specific cells in LNs B cell population. **b,** Representative FACS plots showing the gating strategy to identify the proportion of 9G4 specific B cells among B cell populations. **c**, Representative FACS plots showing the detection of Ro60 specific B cells with tetramers. **d**, Representative FACS plots showing the detection of Spike specific B cells with tetramers. **e**, Analysis of Spike positive B cells in LN showed by a representative flow gating strategy, B cell subsets proportions and comparison of Spike-reactive B cells in the indicated populations of activated naïve, non-B_GC_, B_GC_ and B_PC_. Mann Whitney U test shown for comparisons.

**Extended Data Fig. 5 LN B cells VH mutational state and VH _4-34_ usage. a,** Table showing the mean SHM count for the V genes in the indicated B cell populations for HD and SLE. VH_4-34_ gene is indicated in the red box. **b,** Representative Honeycombs showing HD and SLE B cell clonal distribution (separated by singletons, small clones size 2-5, and large clones >5) and their top-10 ranked-usage of VH genes. **c**, Scatter plots showing the levels of serum 9G4 IgG in the selected donors tested by 10X single cells. Red dots indicate the donors from which VH_4-34_ is ranked in the top-10 used gene, among LN B cells. **d**, Table data showing the 10X donors demographics and serological information, alongside the Top-20 ranked-usage VH_4-34_.

**Extended Data Fig. 6 Testing of VH_4-34_ mAbs and their properties. a,** FACS plots representative of BCB assay with healthy donor Naïve B cells incubated with 5 ug of mAbs. **b,** FACS plots representative of APCB assay with J45.01 Jurkat cell line treated with apoptosis inducer Camptothecin (CPT) and incubated with 5 ug of mAbs the next day. **c**, Scatter box violin plots showing the SHM count levels shown in the mAbs from HD or SLE that bound to Hep-2 for AVY-unmutated and AVY-mutated group of mAbs. Boxes indicate the medians. **d**, Cluster heatmaps showing HD (n=41) and SLE (n=80) tested mAbs in Hep-2 immunostaining and their features of mutation counts as replacement (r) or silent (s) mutations overall in the sequence, or in the CDRH (cdr) or framework (fwr) regions and their sub-regions, alongside their CDR3H properties - obtained from the VDJ repertoire analysis. Mean values are z-score transformed prior to plotting. Kruskal-Wallis’s statistics indicated when significant among the four IFA Hep-2 immunostaining groups (negative, weak, positive, and strong positive).

**Extended Data Fig.7 Properties of mAbs tested for cross-/poly-reactivity**. Table showing the properties and origins of mAbs tested in this studies for all ELISA tests.

## Methods

### Human participants

This research was approved by the Emory University Institutional Review Board (Emory IRB nos. IRB000058515 and IRB00003799) and performed in accordance with all relevant guidelines and regulations. Blood draws and LN-FNA biopsy were obtained after acquiring written informed consent from the participants. Participants were compensated for their time and interest in the study. Healthy individuals (n = 34) and patients diagnosed with SLE (n = 59) were recruited from Emory University Hospital, Emory University Hospital Midtown and Emory/Grady (all in Atlanta in the United States). The demographics of the HD and SLE cohorts and the medications used by patients with SLE are listed in Supplementary Table 1. All donors are de-identified in the figures. Study data were collected, de-identified and managed using the REDCap electronic data capture tool (a secure web platform for building and managing online databases and surveys) hosted at Emory University.

### LN-FNA biopsy and LNMC isolation

Subjects underwent ultra-sound guided axillary lymph node Fine-Needle Aspirate (FN-LNA) biopsy executed by a board-certified Radiologist. Lymph Node Mononuclear Cells (LNMC) were isolated from the cellular suspension by centrifugation at 800g for 5 minutes, and one or two ACK lysis steps. Viability was counted using an automated hemocytometer. Frozen cells were stored at −80C in FBS/10% DMSO.

### PBMC and serum and plasma collection

Peripheral blood samples were collected in green-top (Vacutainer) sodium heparin tubes. Red-top tubes (Vacutainer) were centrifuged for 10 minutes at 800g to collect the serum. Undiluted plasma was collected after centrifugation of the blood for 10 min at 500g and cleared by spun down at 2500g for 15 minutes. Serum and plasma aliquots were stored at −20C. Next, blood samples were diluted (1:2) with PBS, and PBMCs were isolated by Ficoll density gradient centrifugation at 1,000g for 10 minutes. PBMCs were washed twice with R10 complete medium and lysed with ACK lysis buffer. Viability was counted using an automated hemocytometer. Frozen cells were stored at −80C in FBS/10% DMSO. Metadata from all enrolled donors were kept de-identified and used for analysis in this study.

### Flow cytometry

PBMCs or LNMCs stored in frozen vials were gently thawed for 5-10min in the water bath (at 37C), washed and centrifuged at 800g for 5 min and then resuspended in warm R10 complete medium.

A mixture of diluted antibodies was prepared in Super Bright Complete Staining Buffer and added to the samples in a 96-well round-bottom plate at a final volume of 50 μl per well. After each step, antibody staining was blocked with a wash in staining buffer (MACS Buffer, ∼100 μl) and centrifugation of the plate at 800g for 5 min.

For the B cell immunophenotype, after incubation with the tetramers for 1 hour at 4 degrees Celsius, cells were washed and stained with a mixture of anti-immunoglobulins (anti-IgD + anti-IgM + anti-IgA + anti-IgG) for ∼15 min at room temperature (RT) in the dark. The cells were then incubated with the cocktails of the remaining antibodies for ∼40 min at RT in the dark. After washing and centrifugation, cells were stained with L/D Blue (diluted at 1:500 in PBS) for 10 min at RT in the dark. Similarly, for the intracellular staining, samples were stained with the antibody mixture for ∼40 min at RT in the dark, and viable cells were detected with L/D Zombie NIR. Cells were resuspended in ∼180 μl staining buffer, kept on ice and immediately analyzed on a 5L Cytek Aurora flow cytometer using Cytek SpectroFlo software. Flow cytometry data were analyzed using FlowJo v10.10.1.

### Detection of probe- and tetramer-binding antigen-positive B cells

Antigen-specific B cells were detected using tetramer probes or the rat-anti-Idiotype 9G4 antibody. Biotinylated Spike (CAT#AVI10549-050) and RBD (CAT#AVI10500-050) proteins (R&D Systems) or Ro60 protein (rec. human Ro60, Prospect CAT#PRO-329) were multimerized with fluorescently labeled streptavidin (SA) for 1 hour at 4 degrees Celsius. Full-length spike protein was mixed with SA-BV421 using ∼100 ng spike and ∼20 ng SA for each sample (∼4:1 molar ratio). RBD was mixed with SA-AF647 using ∼25 ng RBD and ∼12.5 ng SA (∼4:1 molar ratio). SA-PeCy5.5 was used as a decoy probe. After 1 hour, all tetramers were mixed in free d-biotin (5 μM, Avidity) to minimize cross-reactivity. PBMCs were incubated for 1 hour at 4 degrees Celsius with the mixture of tetramers, washed and stained with surface markers, and labeled for viability with L/D Blue (1:500) for ∼10 minutes before being acquired with the 5L Aurora (Cytek).

### BCB and APCB assay

Monoclonal antibodies were tested at a concentration of 5 μg for cellular binding to naïve B cells (B cell Binding, BCB) or apoptotic cellular debris (Apoptotic cell binding, APCB) as previously reported^9, 23^. Briefly, for BCB assay, total PBMC or isolated B cells from a healthy donor were isolated, FC blocked for 10 minutes, and incubated with each mAb for 30 minutes at 4 degrees Celsius. Cells were then washed, and stained with extracellular antibody staining for 30 minutes at room temperature. After wash, cells were fixed with 4% PFA and acquired on the 5L Aurora (Cytek). For the APCB assay, The CD45-negative Jurkat cell line (J45.1; American Type Culture Collection) was cultured at 1 × 10^6^ cells/ml with 20 μg/ml Camptothecin for ∼18-22 hours to induce apoptosis. A total of 1 × 10^6^ cells was resuspended in 50 μl of MACS buffer in 96-well plate and incubated with each mAb at 5 μg for 30 minutes at 4 degrees Celsius. Cells were washed twice and subsequently stained with extracellular surface staining for 30 minutes at 4 degrees Celsius. After wash, each well was resuspended in ∼140 ul of Annexin V buffer with anti–Annexin V PE and 7-AAD antibodies (used at 2 μl/sample). Each experiment was carried out with at least a negative well control (cells incubated without mAb) and a positive well control (mAb with confirmed BCB or APCB activity).

### Carbodiimide coupling of microspheres to SARS-CoV-2 antigens and Luminex proteomic assays for measurement of anti-antigen antibody

This analysis was carried out as previously described. Briefly, five SARS-CoV-2 proteins were coupled to MagPlex microspheres of different regions (Luminex). The nucleocapsid Wa.1 strain (cat. no. Z03480, expressed in Escherichia coli), nucleocapsid Omicron BA1 strain (cat. no. Z03731, expressed in Escherichia coli), and S1-RBD Wa.1 strain (cat. no. Z03483, expressed in human cells) proteins were purchased from GenScript. The S1 domain Wa.1 strain (cat. no. S1N-C52H2, expressed in HEK293 cells), S2 domain Wa.1 strain (cat. no. S2N-C52H2, expressed in HEK293 cells), S1 domain Omicron BA4.5 strain (cat. no. S1N-C52Hy, expressed in HEK293 cells), and S1-RBD Omicron BA4.5 strain (cat. no. SPD-C522r, expressed in HEK293 cells) proteins were purchased from ACROBiosystems. Each protein was expressed with an N-terminal His6-tag to facilitate purification (>85% purity) and appeared as a predominant single band on SDS–PAGE analysis. Coupling was carried out at 22C following standard carbodiimide coupling procedures. The concentrations of coupled microspheres were confirmed using a Bio-Rad T20 cell counter. Approximately 50 μl of the mixture of coupled microspheres was added to each well of 96-well clear-bottom black polystyrene microplates (Greiner Bio-One) at a concentration of 1,000 microspheres per region per well. All wash steps and dilutions were accomplished using 1% BSA/1X PBS assay buffer. Serum samples were assayed at a 1:500 dilution and surveyed for antibodies to the nucleocapsid (Wa.1 strain), nucleocapsid (Omicron BA1 strain), S1 (Wa.1 strain), S1 (Omicron BA4.5 strain), S2 (Wa.1 strain), RBD (Wa.1 strain), and RBD (Omicron BA4.5 strain) proteins. After incubation for 1 h in the dark on a plate shaker at 800 rpm, the wells were washed five times with 100 μl assay buffer using a BioTek 405 TS plate washer and then applied with 3 μg ml−1 PE-conjugated goat anti-human IgG (Southern Biotech). After 30 min of incubation at 800 rpm in the dark, the wells were washed three times with 100 μl assay buffer, resuspended in 100 μl assay buffer and analyzed using a Luminex FLEXMAP 3D instrument (Luminex) running xPONENT 4.3 software at an enhanced photomultiplier tube setting. The MFI of the anti-IgG detection antibody was measured using the Luminex xPONENT software. The background value of the assay buffer was subtracted from each serum sample result to obtain the net MFI (MFI minus background) value.

### Anti-self and anti-viral antibodies detection with ELISA and LIPS assays

ELISAs were performed *in house*, coating COSTAR 96 well Assay Plate (Corning, CAT#9018) with 3 ug/ml of each antigen: Ro60 (Prospec, CAT#PRO-329), Ro52 (Surmodics, CAT#A12700), La (Surmodics, CAT#A12800), H3 (Cayman, CAT# 10263), LPS (Invitrogen, CAT# 00-4967-93), PCNA (Surmodics, CAT#A15400), Spike (R&D, CAT#RP1210721.46/P210721.02), EBNA1 (aback, CATab138345), SmD3 (Arotec Diagnostics, CAT#S02-01) in PBS overnight at +4C. For dsDNA (Sigma Aldrich, CAT#11691112001), plates were pre-coated with Poly-l-lysin (1:10 diluted with PBS; 0.1% w/v in water) for 30 minutes at 37C, washed and then coated with dsDNA overnight. Plates were blocked with blocking buffer (Blocker Casein in PBS, Thermo Fisher, Cat#375228) for 1 hour at 37C. mAbs were diluted in PBS + 0.2% casein and tested at a concentration of 5 ug. Samples were incubated for 2 hours at room temperatures. Plates were washed 4 times with PBST and 3 times with PBS, and the anti-human IgG HRP secondary antibody (Sigma, Cat# A6029, diluted as 1:5000 in PBS + 0.2% casein buffe) was added for 1 hour at room temperature. Plates were washed as before (a total of 7 times) and each well added with 80 ul of TBM substrate (Thermo Fisher, Cat#002023). Plates were incubated in the dark, and signal was developed for about 10-25 minutes. Signals were stopped with 80 ul of Stop Solution (Invitrogen, cat#SS04). Optical density was read at 450nm with an ELISA Reader (BioTek Synergy 2). 75G12 mAb was used in each ELISAs as a positive control of auto- and poly-reactivity.

LIPS was performed *in-house* to test for Ro60 and Smith D3 positivity using respective luciferase-tagged antigens produced in mammalian COS-1 cells. Ruc-tagged recombinant protein based on light units was first incubated with each serum or plasma sample typically for 1 hour. The reaction mixture was then transferred for an additional hour to a filter plate containing antibody capturing A/G beads (Thermo Scientific CAT#53133. Following the incubation the relative amount of antibody bound was determined measuring the light produced when adding Renilla Luciferase Substrate (Promega, E289B) diluted 1:100 in Renilla Assay Buffer (Promega, E290B). The light units were measured in a Berthold LB 960 Centro luminometer.HEp-2 cell immunostaining

### HEp-2 cell immunostaining

The mAbs were tested at 5 ug using the NOVA Lite Hep-2 Kit (INOVA Diagnostic, Cat#708101), following the manufacturer’s instructions. Briefly, samples were diluted in PBS and added to each well (∼25ul). Slides were incubated in most chambers for 30 min, at room temperature. Slides were gently washed with PBS and incubated for 5 minutes. Secondary anti-human IgG HRP was added to each well, and incubated for 30 minutes and room temperature. Negative (Kit vial) and positive controls (Kit vial and/or SLE positive sera) were added to each slide. Next, slides were washed in PBS for 5 minutes and mounted with mounting medium. Slides with cover slips were acquired on an EVOS M500 Microscope at 10X, 20X, and 40X magnification. Positive/negative signals and patter IFA staining (e.g., Cytoplasmic (C), Homogeneous (H), Speckled (S), Nuclear Rim (NR)) were assigned for each mAb tested.

### SPR analysis of antibody binding kinetics to Spike SARS-CoV-2 and Ro60 protein

The steady-state equilibrium binding of post-SARS-CoV-2-vaccinated human polyclonal samples was monitored at 25 ÅãC using a ProteOn SPR system (Bio-Rad). The purified recombinant protein was captured to a GLC sensor chip with 300 resonance units (RU) in the test flow channels. Serial dilutions (10-, 50- and 250-fold) of freshly prepared samples in BSA–PBST buffer (PBS pH 7.4 with Tween 20 and BSA) were injected at a flow rate of 50ul min−1 (contact duration 240 s) for association, and disassociation was performed over a 1,200 s interval. Responses from the protein surface were corrected for the response from a mock surface and for responses from a buffer-only injection.

All SPR experiments were performed twice. In these optimized SPR conditions, the variation for each sample in duplicate SPR runs was <7%. Antibody off-rate constants, which describe the stability of the antigen–antibody complex (that is, the fraction of complexes that decays per second in the dissociation phase), were determined directly from the interaction of human polyclonal samples with recombinant purified protein using SPR in the dissociation phase only for sensorgrams with maximum RU in the range of 10–150 RU and calculated using the Bio-Rad ProteOn manager software for the heterogeneous sample model as previously described. Off-rate constants were determined from two independent SPR runs.

### Single-cell library preparation, sequencing, and data preprocessing

LNMC suspensions were stained for 10 min with Fc block at +4 degrees Celsius and subsequently stained with hast-tagged tetramer probes. Whole cell suspension prepared as 20,000-30,000 cells per reaction was loaded in each well of the chip. Two independent 10x Genomics Chromium Single Cell Immune Profiling experiments were performed: Exp_003 (SLE, n = 6; HD, n = 2) using Chromium Next GEM Single Cell 5′ Reagent Kits (v2, Dual Index), and Exp_005 (SLE, n = 2; HD, n = 4) using Chromium Next GEM Single Cell 5′ Reagent Kits (v3). Libraries were sequenced on a NovaSeq X Plus system (Illumina) at Novogene and NYU Langone, with sequencing depths of 20,000 reads per cell for gene expression and 5,000 reads per cell for V(D)J libraries. Raw sequencing data were processed with Cell Ranger pipelines (v8.0.0 for Exp_003 and v9.0.0 for Exp_005) to demultiplex, align, and quantify gene expression (GEX) and VDJ reads against the GRCh38 human reference genome.

### Single-cell RNA-seq data analysis

Seurat (v5.3.0) was used to perform quality control, normalization, and subsequent analysis of scRNA-seq data. Initial quality control was performed separately for each sample to exclude low-quality cells with ≥10% mitochondrial gene content or ≤200 detected features. All immunoglobulin genes were subsequently removed from the main gene expression assay to avoid bias from highly expressed antibody transcripts during downstream analyses. Gene expression counts were normalized using the regularized negative binomial regression (SCTransform). Following individual processing of the datasets from the two scRNA-seq experiments, Exp_003 and Exp_005 were integrated and batch-corrected using Harmony (REF). Clusters were then identified by constructing a shared nearest neighbor (SNN) graph on the Harmony-corrected PCA embeddings using ‘FindNeighbors’, followed by clustering with ‘FindClusters’ (Louvain algorithm). Subsequent dimensionality reduction and visualization were performed using uniform manifold approximation and projection (UMAP). Clusters were manually annotated based on core marker genes and differentially expressed genes identified using ‘FindAllMarkers’ function. Major immune cell populations were first defined to identify and separate B cells from other immune cell types. B cells were then subset and re-clustered to delineate the main B cell populations (B_GC_, non-B_GC_, and B_PC_). Each of these populations was subsequently re-clustered to identify and annotate finer subclusters within each group.

### Single-cell BCR repertoire analysis

Cell Ranger VDJ filtered outputs were processed using the Immcantation framework with Change-O to assign V(D)J genes with IgBLAST via ‘AssignGenes.py’ and generate AIRR-compliant rearrangement data. Only productive sequences were retained for downstream analysis, and inferred germline sequences were reconstructed per clone using the Dowser ‘createGermlines’ function with IMGT reference germline sequences. Clonal annotation was performed using SCOPer’s ‘hierarchicalClones’ function. The nearest-neighbor distance threshold for clonal grouping was estimated using SHazaM and applied to define clones based on IGHV and IGHJ gene usage and junction sequence similarity, using a distance threshold of 0.15. Somatic hypermutation (SHM) was quantified using the ‘observedMutations’ function by comparing sequence alignments to inferred germline sequences. Mutation counts and frequencies were calculated using IMGT-defined V-region partitions (IMGT_V_BY_REGIONS), including CDR1, CDR2, FWR1, FWR2, and FWR3. Honeycomb plots were created using the Enclone tool (v0.5.219; 10x Genomics). IgPhyML was used to infer genealogical trees for the selected clones via the Dowser ‘getTrees’ function. Clonal diversity was assessed using Shannon entropy and the Gini coefficient after subsampling each sample to 500 cells to control for differences in the number of recovered cells across samples. Clonal connectivity between B cell subsets was visualized using circos plots generated with the Circlize R package. Networks were constructed based on shared clone identifiers across subsets. Each clone was annotated with a dominant immunoglobulin isotype, defined as the isotype representing ≥75% of sequences within the clone; clones not meeting this threshold were classified as mixed.

### Selection and expression of recombinant mAbs

Variable heavy and light chain sequences were selected based on (i) being a member of VH4-34 family, (ii) being assigned to B cell clusters of naïve, non-B_GC_, B_GC_ or B_PC_, (iii) low versus high mutated clones.

For recombinant expression, mAb were either produced with Twist or in-house in Dr. King laboratory.

For Twist orders, the heavy and light-chain variable sequences were codon optimized, synthesized, and cloned into IgG1 or IgG4 heavy chain constant region or κ _or λ _light chain constant regions, respectively.

For laboratory produced mAb, PCR IgVH and IgVK/L amplicons, generated from the single cell cDNA, were cloned into expression vectors containing the constant regions of human IgG1, for IGHVs or IGK, or IGL for light chain variable regions. Plasmids were sequenced on the Oxford nanopore Minion platform using indexing array barcodes. IgH and IgL plasmids were co-transfected using polyethyleneimine into 293FreeStyle cells in Freestyle expression media (Invitrogen). Additional mab re-production from glycerol stocks or plasmids were carried in the Sanz lab. Briefly, Midiprep plasmids for VH and VL chains were co-transfected into Expi293 cells using FectoPro (Polyplus, Cat#101000014). Supernatants were collected after five days, and mAbs were purified with AmMag Protein A magnetic beads (Genscript). Antibody concentrations were measured with a nanodrop spectrophotometer.

### Statistical analysis

Statistical analysis was carried out using Prism v.9.5.1, v.10.2.2, and v10.0.3 (GraphPad Software) or R coding system with the support of Claude. For each experiment, the type of statistical testing, summary statistics, and levels of significance can be found in the figures and corresponding legends. Levels of significance are indicated as follows: *P < 0.05, **P < 0.01, ***P < 0.001 and ****P < 0.0001.

## Data availability

The flow cytometry dataset and single cell dataset are available upon request from the corresponding authors.

### Acknowledgements

We thank all the donors for their generosity in participating in this research and making this study possible. We thank the laboratory clinical coordinators’ team, as well as the nurses, staff and providers in the Emory Clinics and Hospitals in Atlanta, GA. We also thank the members of the Sanz laboratories for helpful discussions.

This work was supported by the National Institutes of Health grants U19-AI110483 Emory Autoimmunity Center of Excellence (I.S.), and P01AI125180 grant (I.S, F.E.-H.L.), and the LRA DIA (Distinguished Innovator Award) grant (I.S.). The SPR-based antibody measurement was supported by the Food and Drug Administration’s Perinatal Health Center of Excellence (PHCE) project grants #GCBER005 and #GCBER008 to S.K. The content of this publication does not necessarily reflect the views or policies of the Department of Health and Human Services, nor does mention of trade names, commercial products or organizations imply endorsement by the US government. Opinions, interpretations, conclusions, and recommendations are those of the authors and are not necessarily endorsed by the US government. This article is an informal communication and represents our own best judgment. The material in this manuscript and comments do not bind or obligate the FDA.

## Author contributions

C.E.F. and IS conceived and directed this study. C.E.F. and M.G. performed ELISA, flow cytometry, mAb testing used in the study. M.G.V. performed single-cell analysis and prepared figures. C.E.F. analyzed and compiled the resulting data. R.C.W. performed serum testing against viral antigens. O.P., M.A. and S.K. performed sera affinity test with SPR. V.G., C.D.S., J.M.L. provided critical feedback and support in data analysis and interpretation. A.C., J.B., and R.G.K. produced some of the mAb tested in this study. M.N. performed all lymph node FNA biopsies. F.E.L. and A.K. conducted chart review and identified and screened patient samples for study inclusion. C.E.F. and I.S. wrote the manuscript, with other authors providing editorial support.

## Competing interests

I.S. is a consultant for Kyverna Therapeutics SAB, Merck, Novartis, GSK, and Otsuka. F.E.-H.L. is the founder of MicroB-plex, Inc., and has research grants with Genentech. The other authors declare no competing interests.

**Correspondence** and requests for materials should be addressed to Iñaki Sanz or Caterina E. Faliti.

## Notes

### Author Declarations

This research was approved by the Emory University Institutional Review Board (Emory IRB nos. IRB000058515 and IRB00003799) and performed in accordance with all relevant guidelines and regulations.

## References

1. Sanz, I. Rationale for B cell targeting in SLE. Semin Immunopathol 36, 365–375 (2014).

2. Chen, W., et al. SLE Antibody-Secreting Cells Are Characterized by Enhanced Peripheral Maturation and Survival Programs. Res Sq (2023).

3. Tipton, C.M. et al. Diversity, cellular origin and autoreactivity of antibody-secreting cell population expansions in acute systemic lupus erythematosus. Nat Immunol 16, 755–765 (2015).

4. Cappione, A., 3rd et al. Germinal center exclusion of autoreactive B cells is defective in human systemic lupus erythematosus. J Clin Invest 115, 3205–3216 (2005).

5. Mackensen, A. et al. Anti-CD19 CAR T cell therapy for refractory systemic lupus erythematosus. Nat Med 28, 2124–2132 (2022).

6. Müller, F. et al. CD19 CAR-T cells for treatment-refractory autoimmune diseases: the phase 1/2 CASTLE basket trial. Nat Med 32, 1142–1151 (2026).

7. Junt, T. et al. Defining immune reset: achieving sustained remission in autoimmune diseases. Nat Rev Immunol 25, 528–541 (2025).

8. Isenberg, D., Spellerberg, M., Williams, W., Griffiths, M. & Stevenson, F. Identification of the 9G4 idiotope in systemic lupus erythematosus. Br J Rheumatol 32, 876–882 (1993).

9. Richardson, C. et al. Molecular basis of 9G4 B cell autoreactivity in human systemic lupus erythematosus. J Immunol 191, 4926–4939 (2013).

10. Boccitto, M. & Wolin, S.L. Ro60 and Y RNAs: structure, functions, and roles in autoimmunity. Crit Rev Biochem Mol Biol 54, 133–152 (2019).

11. Arbuckle, M.R. et al. Development of autoantibodies before the clinical onset of systemic lupus erythematosus. N Engl J Med 349, 1526–1533 (2003).

12. McClain, M.T. et al. Early events in lupus humoral autoimmunity suggest initiation through molecular mimicry. Nat Med 11, 85–89 (2005).

13. Reed, J.H., Jackson, J., Christ, D. & Goodnow, C.C. Clonal redemption of autoantibodies by somatic hypermutation away from self-reactivity during human immunization. J Exp Med 213, 1255–1265 (2016).

14. Greiling, T.M. et al. Commensal orthologs of the human autoantigen Ro60 as triggers of autoimmunity in lupus. Sci Transl Med 10 (2018).

15. Hung, T. et al. The Ro60 autoantigen binds endogenous retroelements and regulates inflammatory gene expression. Science 350, 455–459 (2015).

16. Faliti, C.E. et al. Disease-associated B cells and immune endotypes shape adaptive immune responses to SARS-CoV-2 mRNA vaccination in human SLE. Nature Immunology (2024).

17. Sasaki, T., et al. Clonal relationships between Tph and Tfh cells in patients with SLE and in murine lupus. bioRxiv (2025).

18. Bocharnikov, A.V., et al. PD-1hiCXCR5- T peripheral helper cells promote B cell responses in lupus via MAF and IL-21. JCI Insight 4 (2019).

19. Thompson, H.L., Smithey, M.J., Surh, C.D. & Nikolich-Žugich, J. Functional and Homeostatic Impact of Age-Related Changes in Lymph Node Stroma. Front Immunol 8, 706 (2017).

20. Foster, W.S., Marcial-Juárez, E. & Linterman, M.A. The cellular factors that impair the germinal center in advanced age. The Journal of Immunology 214, 862–871 (2025).

21. Jaffe, D.B., et al. enclone: precision clonotyping and analysis of immune receptors. bioRxiv, 2022.2004.2021.489084 (2022).

22. Faliti, C.E. et al. Disease-associated B cells and immune endotypes shape adaptive immune responses to SARS-CoV-2 mRNA vaccination in human SLE. Nat Immunol 26, 131–145 (2025).

23. Jenks, S.A. et al. 9G4+ autoantibodies are an important source of apoptotic cell reactivity associated with high levels of disease activity in systemic lupus erythematosus. Arthritis Rheum 65, 3165–3175 (2013).

24. Mayer, C.T. et al. The microanatomic segregation of selection by apoptosis in the germinal center. Science 358 (2017).

25. Liu, Y.-J. et al. Germinal center cells express bcl-2 protein after activation by signals which prevent their entry into apoptosis. European Journal of Immunology 21, 1905–1910 (1991).

26. Spellerberg, M., Chapman, C., Hamblin, T. & Stevenson, F. Dual recognition of lipid A and DNA by human antibodies encoded by the VH4-21 gene. A possible link between infection and lupus. Ann N Y Acad Sci 764, 427–432 (1995).

27. Tur, C. et al. Effects of different B-cell-depleting strategies on the lymphatic tissue. Annals of the Rheumatic Diseases 84, 2065–2074 (2025).

28. Cohen, I.J. et al. Chimeric antigen receptor T cells against the IGHV4-34 B cell receptor specifically eliminate neoplastic and autoimmune B cells. Science Translational Medicine 18, eadr9382 (2026).

29. Liang, E.C. et al. Factors associated with long-term outcomes of CD19 CAR T-cell therapy for relapsed/refractory CLL. Blood Adv 7, 6990–7005 (2023).

30. Greiff, V. et al. A bioinformatic framework for immune repertoire diversity profiling enables detection of immunological status. Genome Med 7, 49 (2015).

31. Stone, J.H. et al. Inebilizumab for Treatment of IgG4-Related Disease. N Engl J Med 392, 1168–1177 (2025).

32. Koneczny, I. Update on IgG4-mediated autoimmune diseases: New insights and new family members. Autoimmunity Reviews 19, 102646 (2020).

33. Scharer, C.D. et al. Epigenetic programming underpins B cell dysfunction in human SLE. Nat Immunol 20, 1071–1082 (2019).

34. Jenks, S.A., Cashman, K.S., Woodruff, M.C., Lee, F.E. & Sanz, I. Extrafollicular responses in humans and SLE. Immunol Rev 288, 136–148 (2019).

35. Sanz, I. Human Atypical B Cells. An Overview. Immunol Rev 334, e70058 (2025).

36. Faliti, C.E. et al. P2X7 receptor restrains pathogenic Tfh cell generation in systemic lupus erythematosus. J Exp Med 216, 317–336 (2019).

37. Curran, C.S., Gupta, S., Sanz, I. & Sharon, E. PD-1 immunobiology in systemic lupus erythematosus. J Autoimmun 97, 1–9 (2019).

38. Saggau, C. et al. Autoantigen-specific CD4^+^ T&#xa0;cells acquire an exhausted phenotype and persist in human antigen-specific autoimmune diseases. Immunity 57, 2416–2432.e2418 (2024).

39. Schickel, J.N. et al. Self-reactive VH4-34-expressing IgG B cells recognize commensal bacteria. J Exp Med 214, 1991–2003 (2017).

40. New, J.S. et al. Human anti-glycan reactivity emerges from B cells utilizing private gene rearrangements that are affinity maturated in germinal centers. Immunity 59, 651–665.e655 (2026).

41. Woodruff, M.C. et al. Chronic inflammation, neutrophil activity, and autoreactivity splits long COVID. Nature Communications 14, 4201 (2023).

42. Woodruff, M.C. et al. Dysregulated naive B cells and de novo autoreactivity in severe COVID-19. Nature 611, 139–147 (2022).

43. Younis, S. et al. Epstein-Barr virus reprograms autoreactive B cells as antigen-presenting cells in systemic lupus erythematosus. Sci Transl Med 17, eady0210 (2025).

44. Collison, J. Epstein–Barr virus reactivation linked to SLE. Nature Reviews Rheumatology 15, 450–450 (2019).

