## Supplementary material for "Defective B cell tolerance in SLE lymph nodes underpins VH_4-34_ “clonal damnation” and PD-1⁺TOX⁺ autoreactive B cells expansion": Ext Data

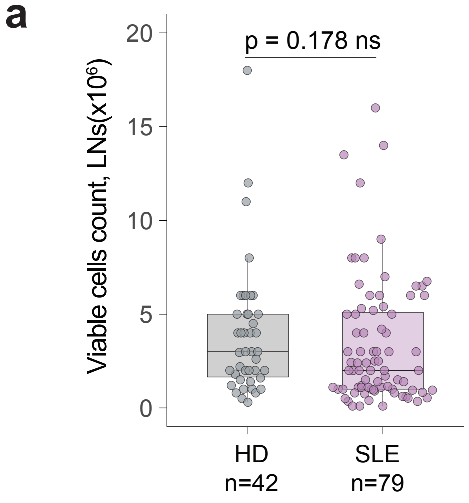

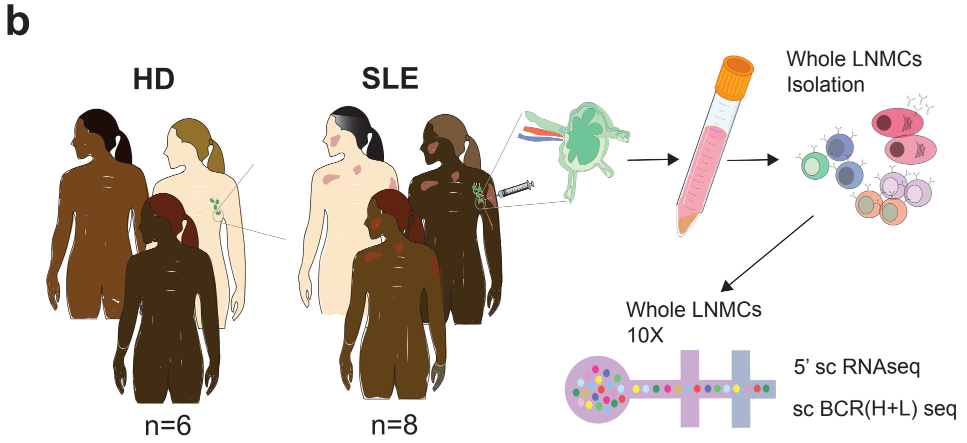

c

Demographics and serology

| 10x_id | exp | group | donor_id | | gender | race | ethnicity | Medication group | SELENA-SLEDAI | years since SLE | Ro60_IgG_AU | ANA_IgG_AU | dsDNA_IgG_IUml | IgG_9G4_ugml |
|---|---|---|---|---|---|---|---|---|---|---|---|---|---|---|
| KF001 | Exp_003 | HD | 7433 | | F | Black | non his | na | na | na | 30.88 | 55.16 | 23 | 89.51 |
| KF002 | Exp_003 | HD | 7529 | | F | Black | non his | na | na | na | 119 | 8.61 | 83.33 | 107.09 |
| KF017 | Exp_005 | HD | 7432 | | F | Black | non his | na | na | na | 5.63 | 30.88 | 84.98 | 98.64 |
| KF018 | Exp_005 | HD | 7439 | | F | Black | non his | na | na | na | 1.93 | 11.37 | 23.6 | 139.08 |
| KF019 | Exp_005 | HD | 7434 | | F | Black | non his | na | na | na | 14.84 | 11.69 | 44.85 | 114.35 |
| KF020 | Exp_005 | HD | 3634 | | F | White | non his | na | na | na | 30.08 | 22.77 | 7.42 | 69.41 |
| KF021 | Exp_005 | SLE | 1539 | | F | White | non his | MMF | 14 | 35 | 8754.98 | 127.25 | 73.85 | 103.79 |
| KF022 | Exp_005 | SLE | 7509 | | F | Black | non his | Saphnelo | 0 | 13 | 89 | 49.77 (*) | 132 | 245.28 |
| KF003 | Exp_003 | SLE | 3894 | | F | Black | non his | BLM | 2 | 7 | 59572.74 | 185.2 | 58 | 334.41 |
| KF004 | Exp_003 | SLE | 1156 | | F | Black | non his | MMF | 4 | 10 | 24141.83 | 196.9 | 3.03 | 328.63 |
| KF005 | Exp_003 | SLE | 7153 | | F | Black | non his | Saphnelo | 2 | 8 | 79413.89 | 136.18 | 910 | 384.1 |
| KF006 | Exp_003 | SLE | 7036 | | F | Black | non his | BLM | 3 | 5 | 62.5 | 156.04 | 138.26 | 84.05 |
| KF007 | Exp_003 | SLE | 254 | | F | Black | non his | MMF | 6 | 20 | 329267.52 | 196.9 | 37.43 | 112.24 |
| KF008 | Exp_003 | SLE | 3293 | | F | Black | non his | BLM | 2 | 7 | 6462.19 | 196.98 | 213.45 | 217.83 |

(*) IFA positive

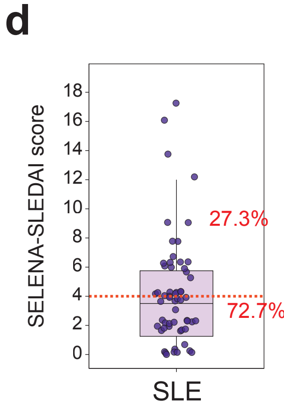

Flow cytometry data

Value: Negative – Positive

| | | | LN-FNA | | | | | | | | | PBMC | | | | | | |
|---|---|---|---|---|---|---|---|---|---|---|---|---|---|---|---|---|---|---|
| 10x_id | exp | group | GC-Tfh | Tefh | CD19⁺ B | $B_{GC}$ | $B_{PC}$ | CD19⁺ ASC | CD19⁻ ASC | non-$B_{GC}$-DN2 | non-$B_{GC}$-DN3 | cTfh | Tph | CD19⁺ B | CD19⁺ ASC | CD19⁻ ASC | $B_{DN2}$ | $B_{DN3}$ |
| KF001 | Exp_003 | HD | 2.19 | 5.85 | 27.5 | 5.89 | 1.27 | 0.77 | 1.01 | 0.51 | 8.25 | 16.7 | 6.94 | 12.4 | 3 | 0.86 | 12.9 | 9.66 |
| KF002 | Exp_003 | HD | 0.59 | 2.87 | 31.1 | 1.4 | 2.08 | 0.88 | 2.61 | 0.26 | 2.63 | 18.3 | 2.61 | 18.2 | 3.18 | 0.07 | 9.79 | 2.01 |
| KF017 | Exp_005 | HD | 0.3 | 2.67 | 25.9 | 1.6 | 0.87 | 0.25 | 1.6 | 0.27 | 9.84 | 4.98 | 0.99 | 10.8 | 0.23 | 0.06 | 1.23 | 14.8 |
| KF018 | Exp_005 | HD | 0.48 | 2.07 | 38.3 | 1.43 | 0.39 | 0.11 | 0.36 | 0.15 | 8.35 | 7.99 | 1.38 | 23.6 | 0.25 | 0.3 | 1.83 | 14.4 |
| KF019 | Exp_005 | HD | 1.01 | 3.9 | 40.6 | 0.76 | 0.9 | 0.5 | 0.55 | 0.34 | 5.37 | 20.5 | 1.72 | 11.9 | 1.37 | 0.07 | 4.69 | 14.3 |
| KF020 | Exp_005 | HD | 1.28 | 2.32 | 44.6 | 2.25 | 0.11 | 0.11 | 0.19 | 0.06 | 3.39 | 7.05 | 1.77 | 17.6 | 1.35 | 0.04 | 3.45 | 8.68 |
| KF021 | Exp_005 | SLE | 0.61 | 2.77 | 12.7 | 1.58 | 1.98 | 0.9 | 0.07 | 0.72 | 3.58 | 4.16 | 2.27 | 11.4 | 0.9 | 0.04 | 3.23 | 13.4 |
| KF022 | Exp_005 | SLE | 17.1 | 12.9 | 58.1 | 35.8 | 3.75 | 2.84 | 4.01 | 1.22 | 15.5 | 11.4 | 10.5 | 28.4 | 3.57 | 0.71 | 7.55 | 6.06 |
| KF003 | Exp_003 | SLE | 1.89 | 12.4 | 13.5 | 10.4 | 6.19 | 3.67 | 17 | 2.42 | 54 | 9.13 | 8.69 | 2.5 | 1.59 | 0.27 | 14.9 | 47.5 |
| KF004 | Exp_003 | SLE | 7.76 | 6.41 | 45.8 | 31.5 | 8.24 | 2.54 | 1.03 | 4.45 | 26.2 | 5.76 | 3.95 | 11.1 | 0.62 | 0.06 | 36.4 | 42.2 |
| KF005 | Exp_003 | SLE | 8.2 | 13.5 | 27.1 | 50.9 | 12.7 | 11 | 1.4 | 17.2 | 15.8 | 7.65 | 5.99 | 4.72 | 10 | 0.05 | 27.1 | 28.4 |
| KF006 | Exp_003 | SLE | 4.38 | 9.42 | 22.1 | 22.1 | 4.37 | 3.66 | 1.02 | 1.16 | 35.9 | 6.85 | 4.82 | 8.6 | 3.63 | 0.67 | 14.3 | 36.2 |
| KF007 | Exp_003 | SLE | 0.97 | 3.96 | 21.8 | 5.68 | 7.78 | 3.2 | 0.5 | 2.99 | 35.5 | 15.6 | 6.59 | 9.79 | 3 | 0.08 | 7.78 | 15.7 |
| KF008 | Exp_003 | SLE | 3.57 | 10.7 | 6.66 | 31.9 | 12.8 | 9.09 | 2.06 | 14.3 | 43.4 | 10.4 | 1.4 | 4.35 | 1.46 | 0.04 | 38.6 | 37.3 |

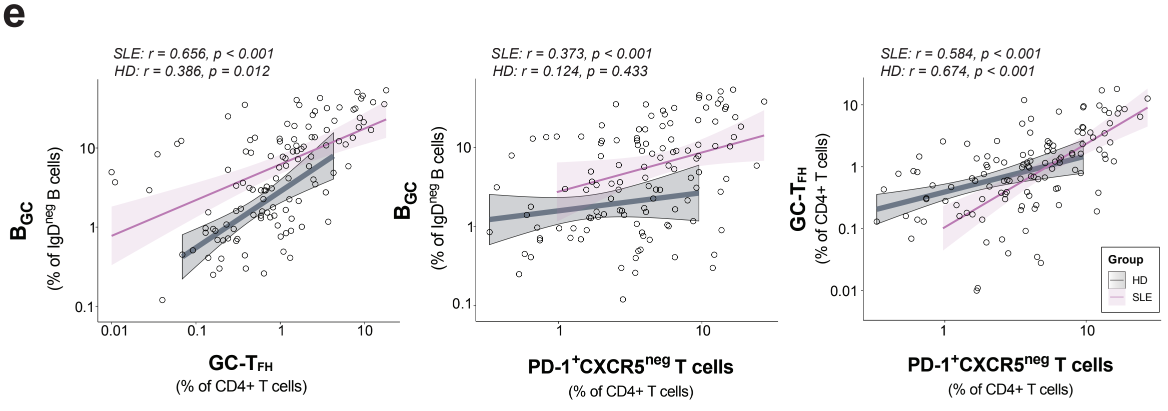

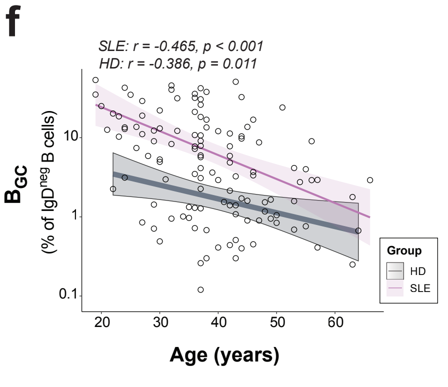

### Extended Data Figure 1

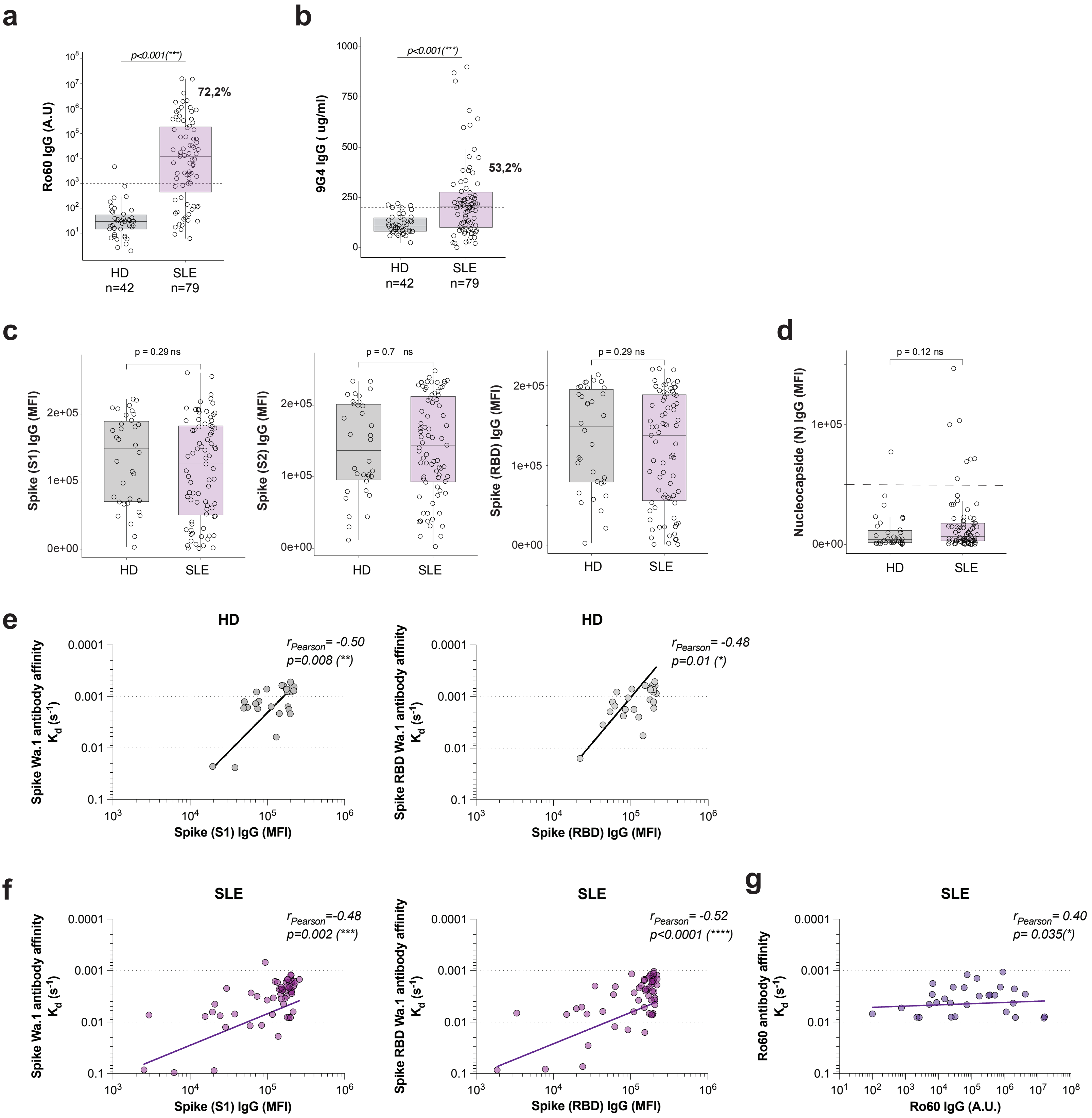

### Extended Data Figure 2

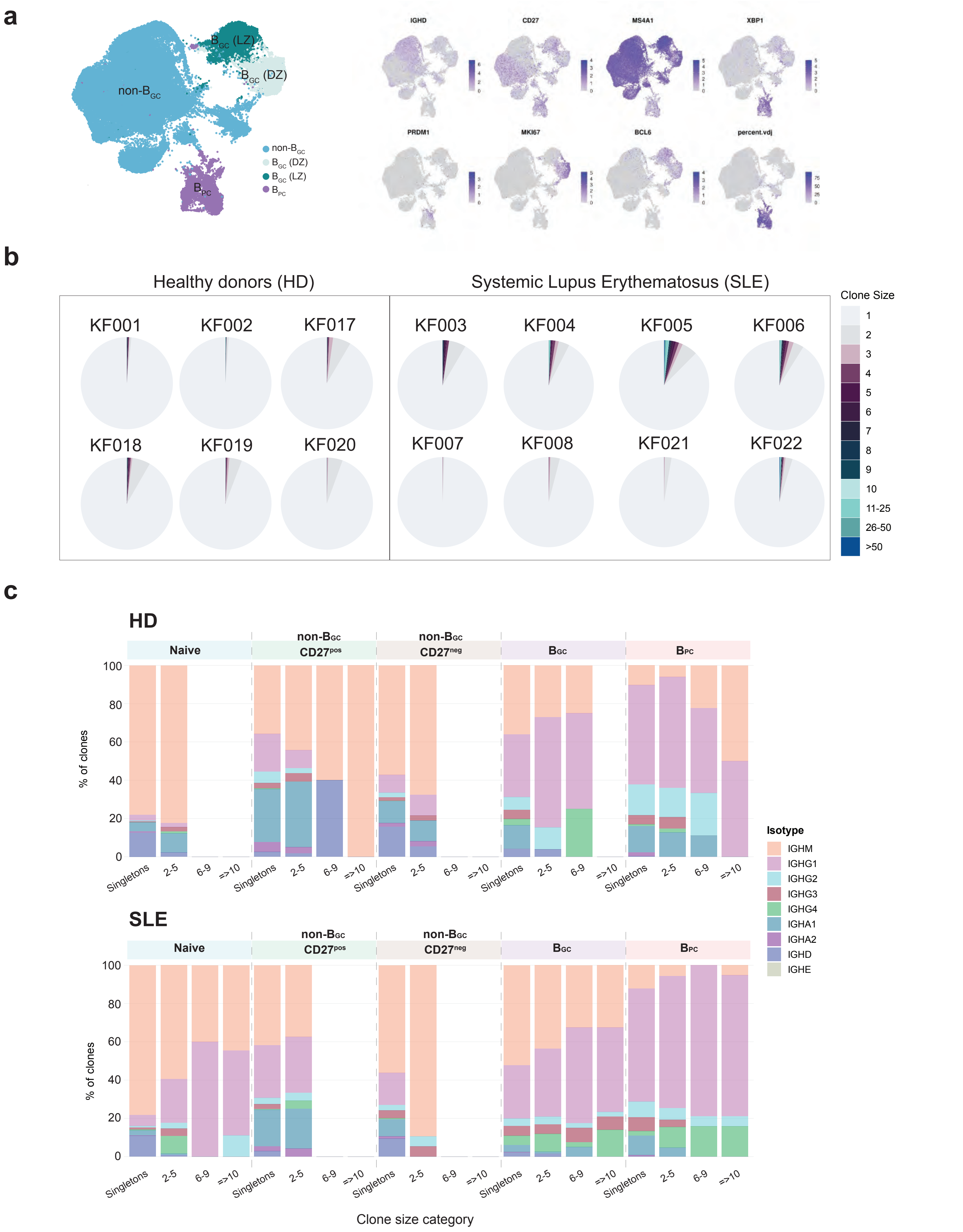

### Extended Data Figure 3

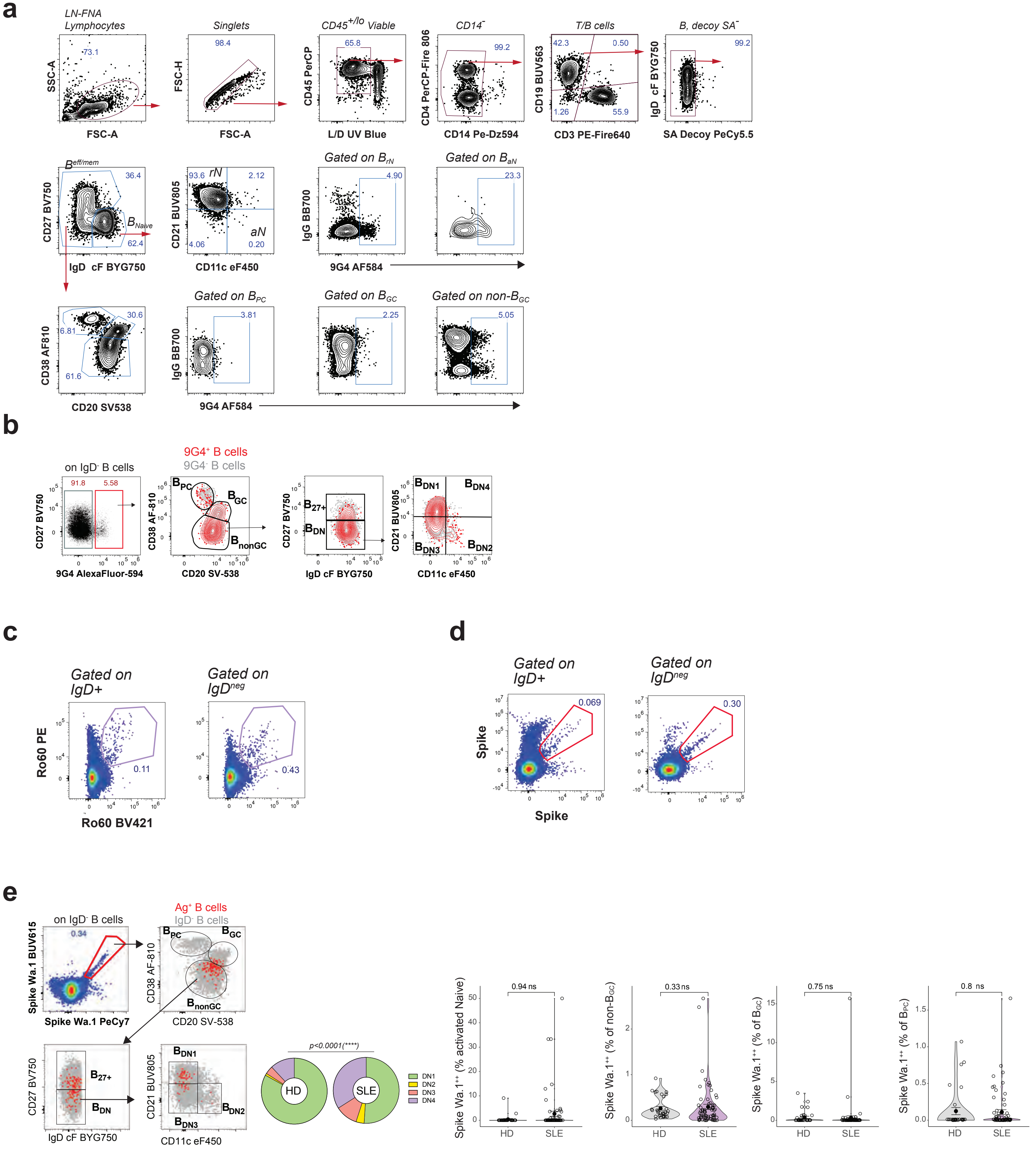

### Extended Data Figure 4

**a**

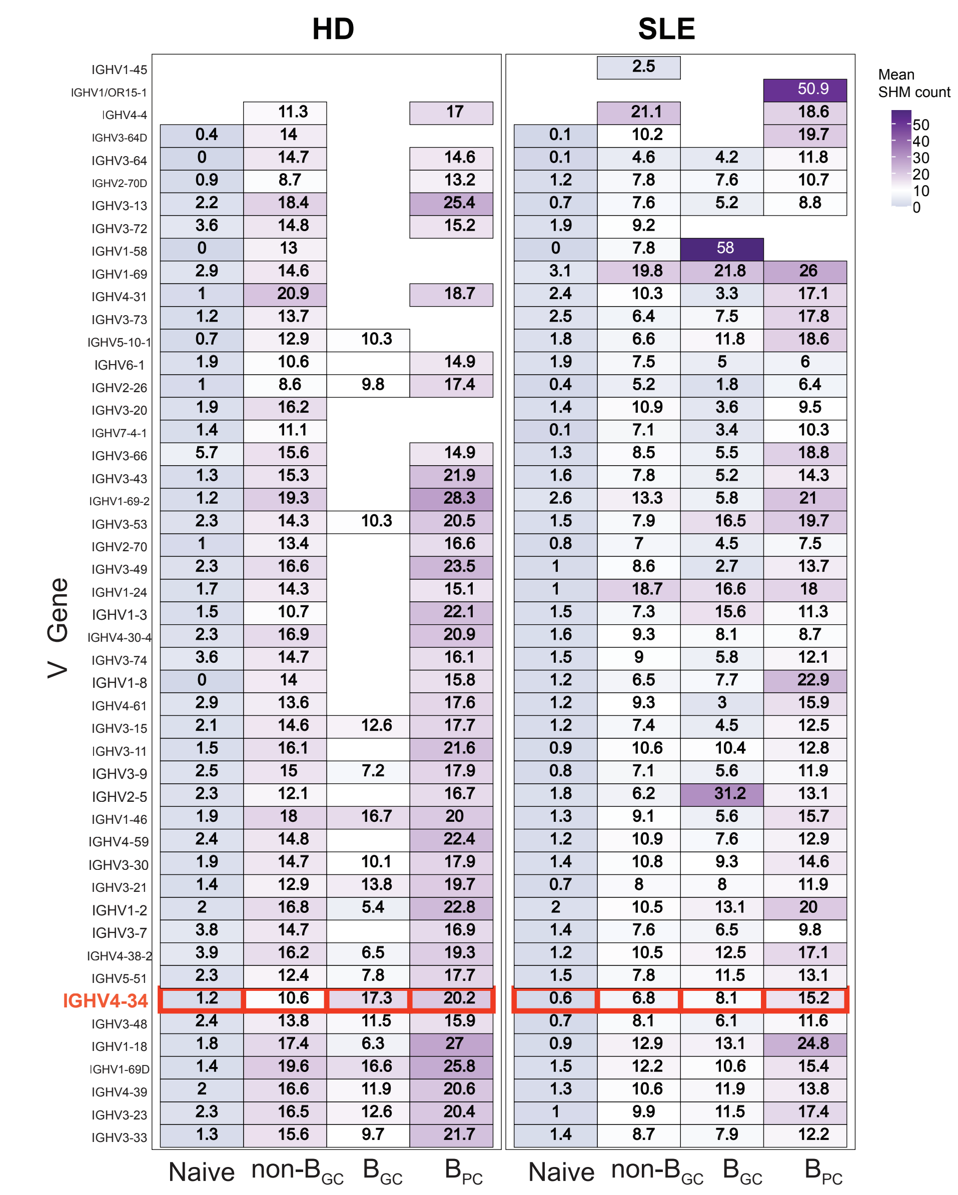

**b**

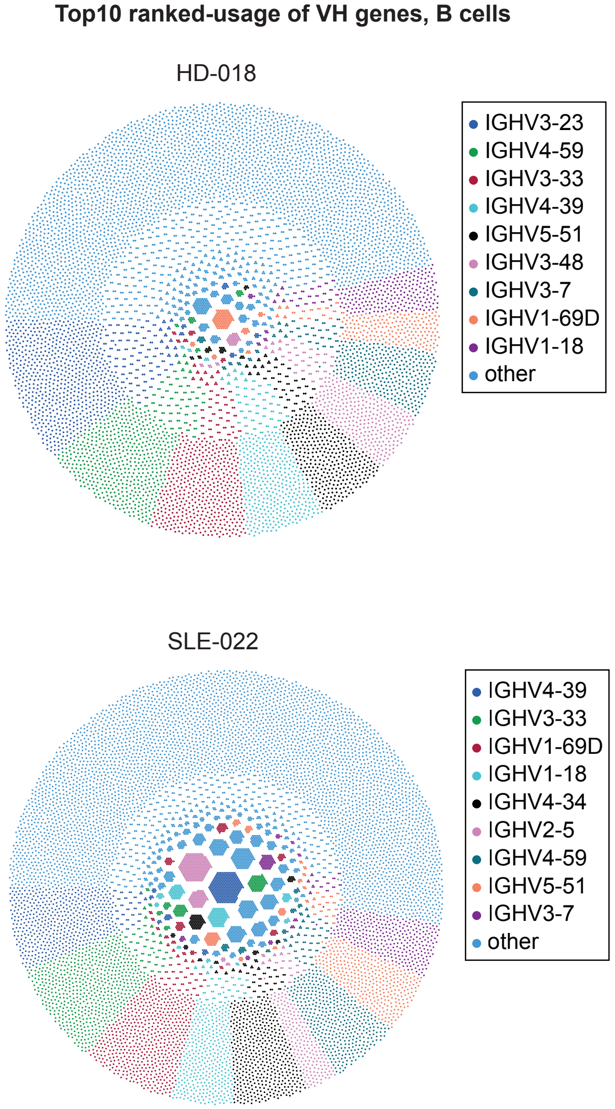

**c**

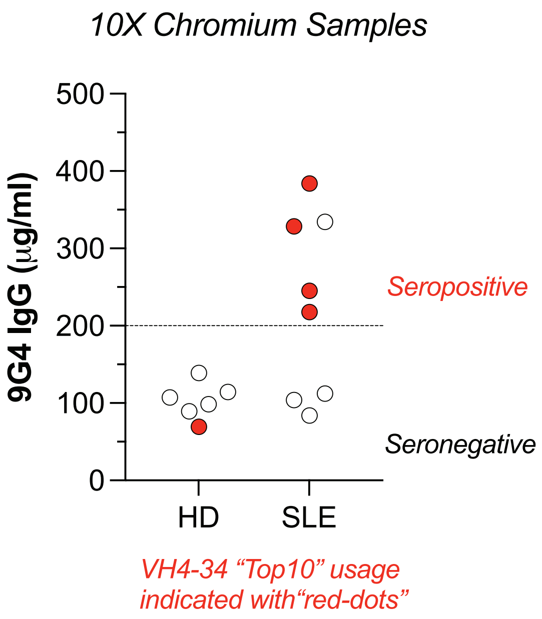

**d**

*10X Chromium Samples*

Demographics and serology

| 10x_id | exp | group | donor_id | gender | race | ethnicity | Medication group | SELENA-SLEDAI | years since SLE | IgG_9G4_ugml | Rank "Top-20" VH4-34 usage |
|---|---|---|---|---|---|---|---|---|---|---|---|
| KF001 | Exp_003 | HD | 7433 | F | Black | non his | na | na | na | 89.51 | 15th |
| KF002 | Exp_003 | HD | 7529 | F | Black | non his | na | na | na | 107.09 | 11th |
| KF017 | Exp_005 | HD | 7432 | F | Black | non his | na | na | na | 98.64 | 11th |
| KF018 | Exp_005 | HD | 7439 | F | Black | non his | na | na | na | 139.08 | 13th |
| KF019 | Exp_005 | HD | 7434 | F | Black | non his | na | na | na | 114.35 | 15th |
| KF020 | Exp_005 | HD | 3634 | F | White | non his | na | na | na | 69.41 | 3rd |
| KF021 | Exp_005 | SLE | 1539 | F | White | non his | MMF | 14 | 35 | 103.79 | 18th |
| KF022 | Exp_005 | SLE | 7509 | F | Black | non his | Saphnelo | 0 | 13 | 245.28 | 5th |
| KF003 | Exp_003 | SLE | 3894 | F | Black | non his | BLM | 2 | 7 | 334.41 | 15th |
| KF004 | Exp_003 | SLE | 1156 | F | Black | non his | MMF | 4 | 10 | 328.63 | 6th |
| KF005 | Exp_003 | SLE | 7153 | F | Black | non his | Saphnelo | 2 | 8 | 384.1 | 8th |
| KF006 | Exp_003 | SLE | 7036 | F | Black | non his | BLM | 3 | 5 | 84.05 | 13th |
| KF007 | Exp_003 | SLE | 254 | F | Black | non his | MMF | 6 | 20 | 112.24 | 15th |
| KF008 | Exp_003 | SLE | 3293 | F | Black | non his | BLM | 2 | 7 | 217.83 | 4th |

Value: Negative — Positive

### Extended Data Figure 5

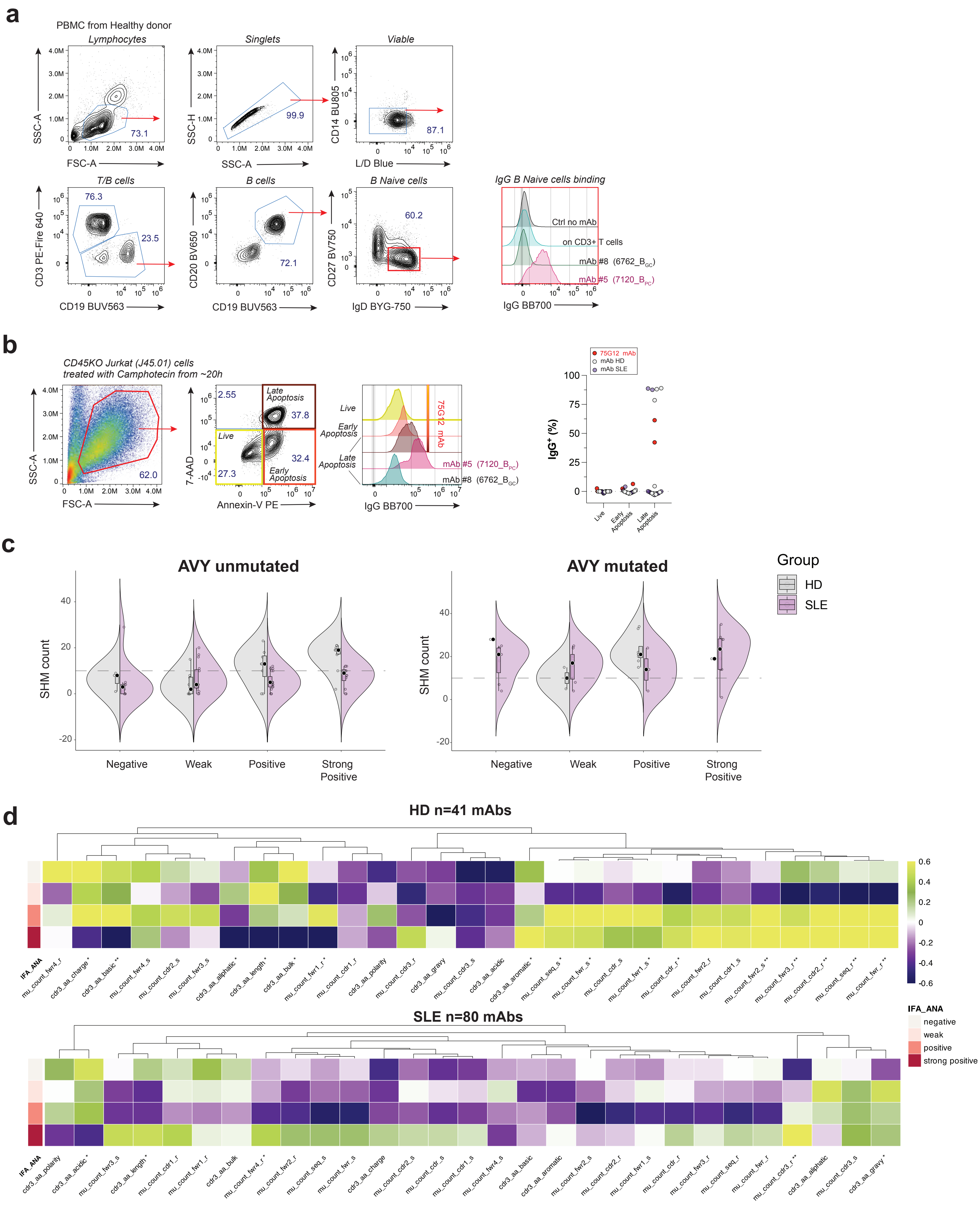

### Extended Data Figure 6

mAbs table

| Group | mAb ID | Donor ID | Clone ID | B cluster | AVY mutated? |
|---|---|---|---|---|---|
| HD | 204 | KF001 | 6596 | C0_BNaive | NO |
| HD | 123 | KF017 | 0_25968 | C0_BNaive | NO |
| HD | 127 | KF020 | 0_25185 | C0_BNaive | NO |
| HD | 30 | KF001 | 6989 | C1_BNaive | NO |
| HD | 126 | KF020 | 0_25152 | C1_BNaive | YES |
| HD | 128 | KF020 | 0_25187 | C1_BNaive | NO |
| HD | 25 | KF001 | 6689 | C2_BCD27+ | YES |
| HD | 28 | KF001 | 6865 | C2_BCD27+ | NO |
| HD | 56 | KF020 | 0_25196 | C2_BCD27+ | NO |
| HD | 38 | KF002 | 6958 | C3_B | NO |
| HD | 36 | KF002 | 6808 | C4_Bact | NO |
| HD | 37 | KF002 | 6852 | C4_Bact | YES |
| HD | 39 | KF002 | 6966 | C4_Bact | NO |
| HD | 205 | KF002 | 7194 | C4_Bact | YES |
| HD | 137 | KF017 | 0_26098 | C4_Bact | NO |
| HD | 124 | KF018 | 0_26019 | C7_B | NO |
| HD | 29 | KF001 | 6986 | DZ | NO |
| HD | 26 | KF001 | 6774 | LZ | NO |
| HD | 27 | KF001 | 6804 | LZ | NO |
| HD | 33 | KF001 | 7099 | LZ | NO |
| HD | 32 | KF001 | 7095 | LZ/Int | YES |
| HD | 34 | KF001 | 7162 | Pre-PB | NO |
| HD | 40 | KF002 | 7029 | PC_0 | NO |
| HD | 567 | KF002 | 6826 | PC_0 | NO |
| HD | 516 | KF019 | 0_25282 | PC_3 | YES |
| HD | 517 | KF019 | 0_25282 | PC_3 | YES |
| HD | 520 | KF019 | 0_25282 | PC_3 | YES |
| HD | 521 | KF019 | 0_25282 | PC_3 | YES |
| SLE | 1 | KF005 | 6948 | C0_BNaive | YES |
| SLE | 16 | KF005 | 6694 | C0_BNaive | YES |
| SLE | 19 | KF006 | 6939 | C0_BNaive | NO |
| SLE | 22 | KF004 | 7007 | C0_BNaive | NO |
| SLE | 24 | KF003 | 7115 | C0_BNaive | YES |
| SLE | 207 | KF004 | 6783 | C0_BNaive | YES |
| SLE | 114 | KF004 | 6838 | C0_BNaive | NO |
| SLE | 117 | KF005 | 6861 | C0_BNaive | YES |
| SLE | 118 | KF005 | 6861 | C0_BNaive | NO |
| SLE | 119 | KF005 | 6861 | C0_BNaive | NO |
| SLE | 120 | KF006 | 7136 | C0_BNaive | NO |
| SLE | 121 | KF007 | 6708 | C0_BNaive | NO |
| SLE | 129 | KF021 | 0_25873 | C0_BNaive | NO |
| SLE | 133 | KF022 | 0_25941 | C0_BNaive | NO |
| SLE | 134 | KF022 | 0_25920 | C0_BNaive | NO |
| SLE | 136 | KF022 | 0_25985 | C0_BNaive | NO |
| SLE | 17 | KF004 | 6783 | C1_BNaive | NO |
| SLE | 131 | KF022 | 0_25000 | C1_BNaive | YES |
| SLE | 11 | KF005 | 6606 | C2_BCD27+ | YES |
| SLE | 115 | KF005 | 6762 | C2_BCD27+ | YES |
| SLE | 6 | KF005 | 7120 | C3_B | NO |
| SLE | 78 | KF022 | 0_25822 | C4_Bact | NO |
| SLE | 122 | KF008 | 6603 | C7_B | NO |
| SLE | 2 | KF005 | 6948 | DZ | YES |
| SLE | 13 | KF005 | 6645 | DZ | YES |
| SLE | 15 | KF005 | 6694 | DZ | YES |
| SLE | 113 | KF004 | 6783 | DZ | NO |
| SLE | 116 | KF005 | 6861 | DZ | NO |
| SLE | 4 | KF005 | 7120 | DZ/Int | NO |
| SLE | 9 | KF005 | 6606 | LZ | NO |
| SLE | 18 | KF006 | 6939 | LZ | NO |
| SLE | 64 | KF022 | 0_25766 | LZ | NO |
| SLE | 73 | KF022 | 0_25385 | LZ | NO |
| SLE | 211 | KF004 | 6783 | LZ | NO |
| SLE | 130 | KF022 | 0_25558 | LZ | NO |
| SLE | 132 | KF022 | 0_25766 | LZ | YES |
| SLE | 8 | KF005 | 6762 | LZ/Int | NO |
| SLE | 138 | KF005 | 6861 | LZ/Int | NO |
| SLE | 210 | KF004 | 6783 | Pre-PB | NO |
| SLE | 135 | KF022 | 0_26027 | PC_0 | NO |
| SLE | 10 | KF005 | 6606 | PC_1 | YES |
| SLE | 12 | KF005 | 6645 | PC_1 | YES |
| SLE | 14 | KF005 | 6645 | PC_1 | YES |
| SLE | 20 | KF005 | 6772 | PC_1 | YES |
| SLE | 209 | KF004 | 6783 | PC_1 | NO |
| SLE | 112 | KF004 | 6783 | PC_1 | NO |
| SLE | 23 | KF003 | 7115 | PC_2 | NO |
| SLE | 3 | KF005 | 6948 | PC_3 | YES |
| SLE | 5 | KF005 | 7120 | PC_3 | NO |
| SLE | 7 | KF005 | 6762 | PC_3 | NO |
| SLE | 21 | KF005 | 6886 | PC_3 | YES |

### Extended Data Figure 7
